# A Computational Ethology Approach for Characterizing Behavioral Dynamics in Bipolar Disorder

**DOI:** 10.1101/2024.11.14.24317348

**Authors:** Zhanqi Zhang, Chi K. Chou, Holden Rosberg, William Perry, Jared W Young, Arpi Minassian, Gal Mishne, Mikio Aoi

**Affiliations:** Department of Computer Science and Engineering, University of California San Diego, La Jolla, CA; Department of Mathematics, La Jolla, CA; Department of Psychiatry, University of California San Diego, La Jolla, CA; Halıcıoğlu Data Science Institute, University of California San Diego, La Jolla, CA; Department of Electrical and Computer Engineering, University of California San Diego, La Jolla, CA; Department of Neurobiology, University of California San Diego, La Jolla, CA

## Abstract

Recent technologies for quantifying behavior have revolutionized animal studies in social, cognitive, and pharmacological neurosciences. However, comparable studies in understanding human behavior, especially in psychiatry, are lacking. In this study, we utilized data-driven machine learning to analyze natural, spontaneous open-field human behaviors in people with euthymic bipolar disorder (BD) and non-BD participants. Our computational paradigm identified representations of distinct sets of actions (motifs) that capture the physical activities of both groups of participants. We propose novel measures for quantifying dynamics, variability, and stereotypy in BD behaviors. These fine-grained behavioral features reflect patterns of cognitive functions of BD and better predict BD compared with traditional ethological and psychiatric measures and action recognition approaches. This research represents a significant computational advancement in human ethology, enabling the quantification of complex behaviors in real-world conditions and opening new avenues for characterizing neuropsychiatric conditions from behavior.

## Main

Behavior, particularly in novel contexts, can provide valuable insights into neuropsychiatric conditions and illness states. Open field studies, which observe individuals in unstructured environments, have the potential to illuminate how different conditions manifest in real-world settings. Bipolar disorder (BD), a chronic psychiatric illness that can have a devastating impact on quality of life, is associated with hallmark behavioral features such as increased energy, which often manifests as greater motor activity and engagement in goal-directed behaviors^1^. Quantifying such behavior holds promise for identifying symptoms and advancing treatment approaches. Contemporary machine learning technologies offer tools to automate the identification of signature behavior patterns, which may provide a unique lens into the underlying brain functions associated with BD and other neuropsychiatric illnesses.

Clinical assessments of psychiatric disorders still rely heavily upon observer-rating scales such as the Hamilton Depression Rating Scale (HAM-D)^2,3^, and various self-reported rating scales such as Young Mania Rating Scale (YMRS)^4^. While valuable, these tools face well-recognized limitations: observer-rating scales condense multidimensional behavioral experiences into discrete categories that are susceptible to subjective interpretation and interrater variability^5,6^. Similarly, self-reported measures can be influenced by recall bias and may fail to capture the nuances of fine motor behaviors^4,7^. For example, the YMRS term ‘Increased Motor Activity-Energy’ encapsulates symptoms that may also appear in conditions such as attention deficit hyperactivity disorder (ADHD). These scales aggregate behavior across varied contexts and timeframes— such as work, home, and leisure activities, which may not best represent real-time behavior. Thus, a continuous, data-driven quantification of context-specific behavior could enhance the precision of psychiatric assessments.

Behavioral manifestations of BD are further complicated by its cyclical nature, as BD is characterized by recurring episodes that alternate between mania and depression^8^. For example, while individuals experiencing episodes of mania often display observable high motor activity, those in a euthymic (affectively stable) state, defined by the absence of mania, hypomania, or a depressed state, may exhibit behaviors that are only subtly different from those of healthy individuals. Moreover, individual differences in life history, pathology, and symptom expression further complicate the ability to identify consistent patterns. This challenging combination of subtlety and heterogeneity provides the rationale for machine learning-driven analyses, which are capable of identifying subtle and individualized behavioral features.

Some progress has recently been made in the quantification of undirected human behavior in medical settings. For instance, symbolic movement representations learned from continuous wearable device data have demonstrated success in characterizing motor dysfunction in Parkinson’s disease in humans^9^. Similarly, the human Behavioral Pattern Monitor (hBPM), a human version of the classic rodent open-field activity assessment, was developed to better quantify human exploratory behavior^10^. hBPM uses spatial information (Spatial-D; see Methods) and temporal statistics to identify signature patterns of behavior of human patients^10,11^. However, the hBPM still relies on extensive manual labeling of behavior using *a priori* established criteria. This time-consuming process is susceptible to subjective biases in behavioral labels and can be undermined by insufficient inter-rater reliability. Moreover, manual observer-based methods face challenges in scaling to the extensive sizes of modern datasets. To address these challenges, automated, data-driven approaches are necessary to explore behavioral patterns in an unbiased and scalable manner.

Behavior is composed of repeated patterns, or behavioral motifs, that can provide insights into underlying psychological or physiological processes^12–14^. Motifs, which represent recurring, identifiable sequences of actions, reactions, or responses, are exhibited as a characteristic feature of a population. While motifs are often described qualitatively in rating scales, such as HAM-D’s description of “agitation” (i.e., *fidgetiness*; *hand wringing*, or *nail-biting*), they are rarely captured in manual behavior annotations due to their subtlety. This challenge is further compounded by an issue of compositionality: manual annotations primarily focus on broad, coarse actions (e.g., sitting, standing, walking), while finer-grained behaviors like fidgeting often occur concurrently with these larger movements. As a result, these micro-behaviors are either overlooked or collapsed into general action categories, limiting the resolution of behavioral assessments. These omissions raise an important question: can we automatically and quantitively identify motifs from free-moving, spontaneous human behavior in a rich real-world context?

Progress towards this direction has been made in animal behavior analysis, where pose estimation (e.g., DeepLabCut^15^, DeepPoseKit^16^, Deep Graph Pose^17^, DeepOF^18^, and SLEAP^19^) serves as inputs to automated behavioral segmentation methods (e.g., MoSeq-based models^20,21^, VAME^22^, MotionMapper^23–25^, B-SOiD^26^, SimBA^27^, and DBE^28^), which have proven useful for identifying stereotyped behavioral motifs that can be related to neurological^24^ and pharmacological manipulations^14^ in animals. In human, pose estimation technologies (e.g., MoveNet^29^, OpenPose^30^, PoseNet^31^, and MMPose^32^) have demonstrated impressive accuracy in tracking movements, However, both pose-based and video-based human action recognition models (e.g., TSN^33^, I3D^34^, and S3D^35^) are limited by their reliance on predefined action labels, camera angles, and narrow human behavioral scopes, making them less suited for capturing the complexity of behaviors made manifest by psychiatric conditions in the environments in which they are routinely monitored. “Unsupervised” machine learning approaches, in contrast, analyze data without requiring predefined action categories, enabling the discovery of patterns that might reflect individualized or group-level behavioral differences with minimal bias.

In this study, we combined pose tracking with automated behavior motif segmentation to identify spontaneous human behavior in a lab-based real-world context (the hBPM assay). To differentiate individuals with euthymic (affectively stable) BD from healthy comparisons (HC), we developed measures for quantifying behavioral temporal structure, transition dynamics, stereotypy, and expression variability. Using an unsupervised machine learning framework, we identified behavioral features that aligned with clinical observations of BD while also uncovering subtle patterns that may escape human annotation. Our approach demonstrates the potential of computational ethology in clinical psychiatry, offering a scalable, objective method to characterize behavioral markers in neuropsychiatric conditions. These findings have broad implications for enhancing symptoms understanding, tracking symptom fluctuations, and improving treatment strategies through data-driven behavioral phenotyping.

## Results

Study participants have been described previously in hBPM studies^36^. Briefly, 24 participants (10 men) were diagnosed with euthymic (affectively stable) bipolar disorder (BD). Twenty-three were diagnosed with BD Type I or Type II, and one participant was diagnosed with the cyclothymic subtype of BD. All diagnoses were determined by the Structured Clinical Interview for the DSM-IV^37^. All participants gave written consent and were assessed by the YMRS (to assess symptoms of mania) and HAM-D (to assess symptoms of depression). Higher scores on these measures reflect more severe symptoms of mania or depression. All BD participants were in a euthymic state as defined by scores of HAM-D < 10 and YMRS < 12. Healthy comparison (HC) volunteers (n = 24; 12 men) who had never met the DSM-IV criteria for neurological or psychiatric disorders participated in the study as the HC group. Most BD patients were treated with atypical antipsychotics, most commonly Abilify, and mood stabilizers, with Lamictal being the most frequently used. The HC and BD groups were not significantly different on the majority of demographic variables (gender, race, and ethnicity were tested using the Chi-square test, p-value: 0.67, 0.74, 1.00; age, and education level were compared using two-sample t-test yielding p-values: 0.01, 0.87, **Supplementary Table 1**). Given statistically significant differences in age, we included age as a covariate in our analyses to ensure that any observed group differences in behavioral features were not driven by age (**Supplementary Table 5**).

Each participant was introduced to a previously unexplored room containing furniture and small objects along the periphery of the room (**Supplementary Fig. 1**) and remained there for 15 minutes. Videos were recorded from a commercial camera with a fisheye lens placed at the center of the ceiling (**Fig. 1a**). Human experts in previous hBPM studies reviewed the recordings to count occurrences of exploratory action categories, such as “sitting” and “standing.”¹¹ For full details, please refer to **Methods**.

**Fig. 1.**
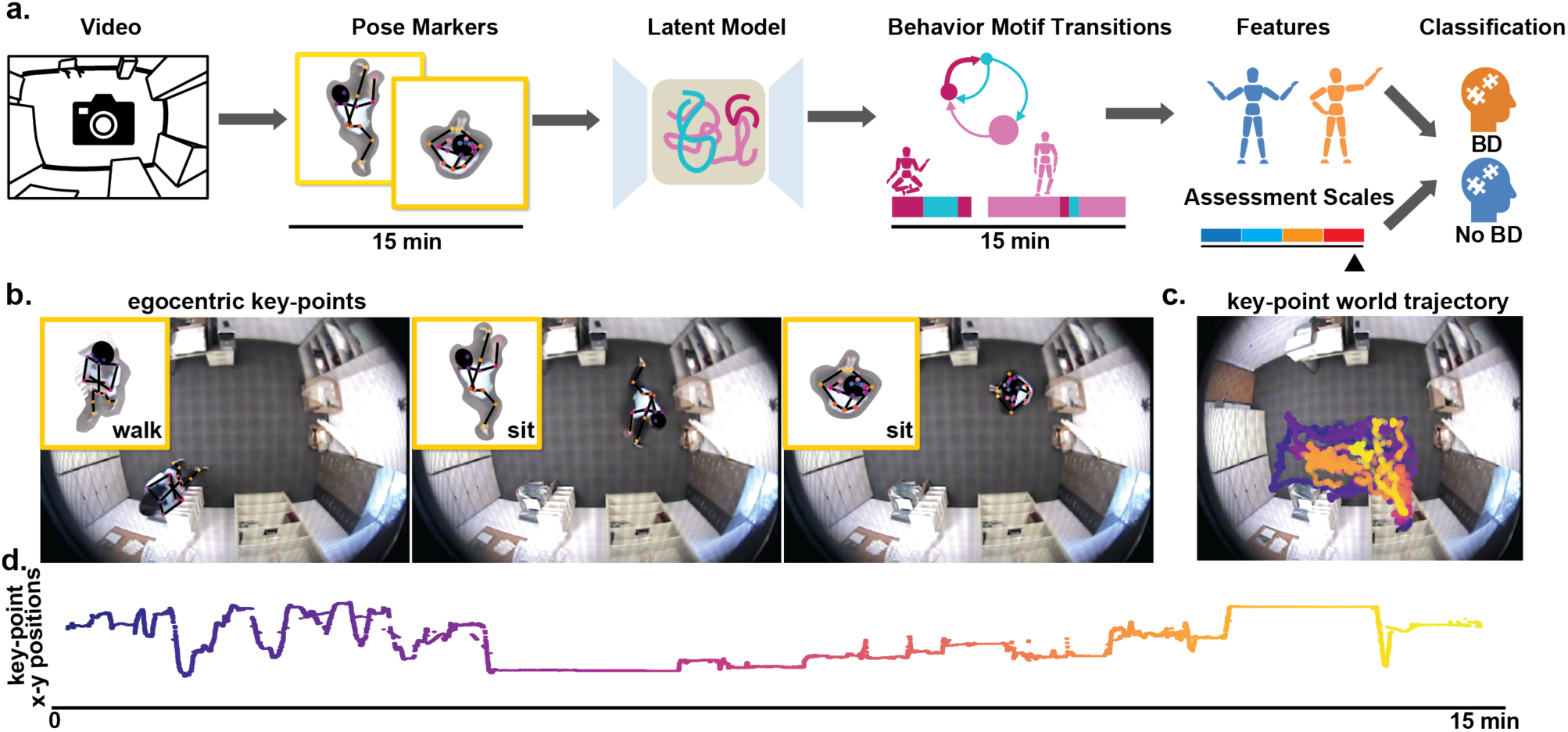
Data and Methods. **a.** Videos of free-moving human behavior from participants with bipolar disorder (BD) during euthymic episodes and healthy comparison (HC) participants for 15 minutes in an unexplored room with objects. We utilized DeepLabCut to label 20 markers placed on key-points of human participants (e.g., elbows). Pose markers were fed into a latent-variable model and the latent representations were used to segment the videos into motifs. We identified hallmark behavioral features that characterized BD in different time scales and these features were used to classify if a participant is from the BD or HC groups. Classification was benchmarked against assessment scales YMRS and HAMD and other action segmentation approaches. **b.** Three example frames from the videos of human behavior with key-points marking the skeleton. Inset: Egocentric view of the human skeleton with key-points are shown with action label from manual behavior annotations. **c.** Example of center-of-feet key-point x-position trajectory in the room. **d.** Trajectory of the center-of-feet key-point x-position over time.

### A Latent-variable model identified context-dependent behavioral motifs of human participants

While the full repertoire of human behaviors is vast, we expect the distribution of behaviors a person expresses in a given context (the hBPM room) to be highly constrained and specific. Therefore, to best characterize the distribution of naturalistic behaviors relevant to the context of our experiment, we required an unbiased way of annotating our video data. Specifically, we developed a data-driven approach with three key functional modules: (1) pose estimation (DeepLabCut) for accurately labeling anatomical key points of the human participants in every frame^15^ (**Fig. 1b-d**), (2) a latent-variable model (VAME) for embedding these key points into a low-dimensional representation^22^ (**Fig. 2a, b**), and (3) clustering on the latent representation to provide a set of behavioral motifs corresponding to distinct actions or sequences of actions, from which we developed metrics to evaluate the behavior dynamics (**Fig. 2c, d**). Here, DeepLabCut was utilized because most off-the-shelf action recognition models are optimized for side or front perspectives and struggle with occlusions and foreshortening inherent in overhead views. To evaluate whether our approach identified patterns of context-dependent, naturalistic human behaviors, we compared these motifs to manually annotated labels determined by clinically trained human experts; as well as pre-trained computer vision (CV) action detection models MMAction^32^ and S3D^35^ which automatically generated a set of labels (**Supplementary Fig. 3a, b**). As an additional control, we applied k-means clustering to the key points themselves (rather than the latent coordinates) to obtain an alternative set of clusters.

**Fig. 2.**
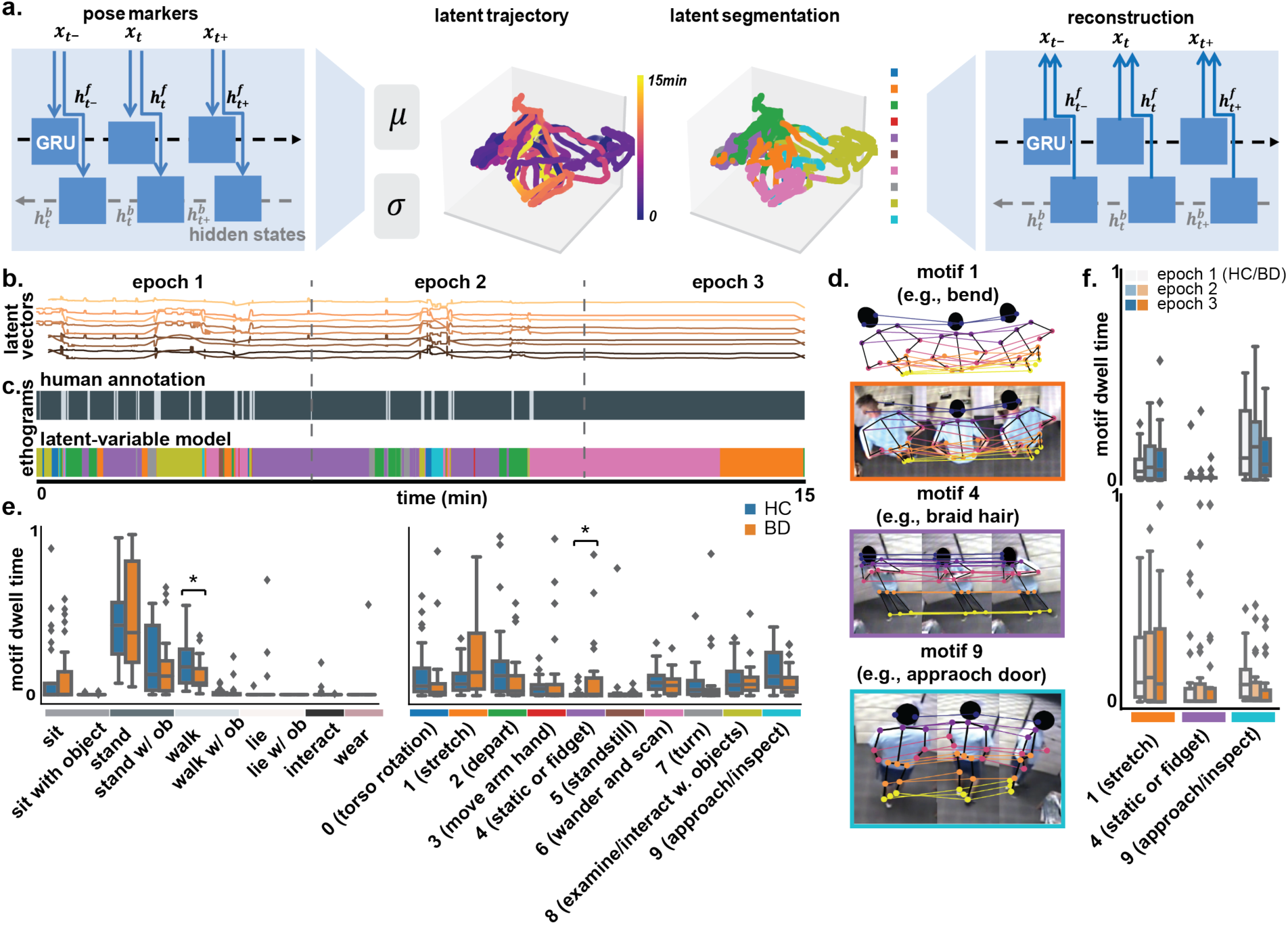
Latent-variable Model and Dwell time. **a.** Pose markers were fed into the VAME variational autoencoder and the latent representations were used to segment motifs. The input were the past *x_t_*_−_, current *x_t_*, and next *x_t_*_+_ pose markers time series which were encoded as corresponding hidden states. The model would learn to reconstruct the input, and the learned latent representation was a 15-min vector that were segmented into motifs. **b.** Example of latent vectors for video in Fig. 1b. **c.** Top: Each video was manually annotated by experts into 10 behavior categories (e.g., sit, stand). Ethogram of manual annotation. Bottom: Ethograms of motif segmentation from latent segmentation. **d.** Human skeletons of motif 1 (upper body bend example); skeletons overlaying on frames of motif 1(*upper body bend* example), motif 4 (*braid hair* example) and motif 9 (*approach door* example) in the dataset. **e.** Motif dwell time (normalized to range [0, 1]) from human annotation (left) and latent variable model (right) in BD (orange) and HC (blue). **f.** Motif dwell time for motif 1, motif 4 and motif 9 in three epochs in BD (light to dark shades of orange), and HC (light to dark shades of blue). Significance marked by asterisks.

We found the distribution of manually labeled behaviors was imbalanced — among 48 videos, the vast majority of time frames are labeled as “stand” or “walk” (median(IQR) BD: 65.2%(34.7%), 17.9%(23.1%); HC: 77.3%(55.3%) 7.9%(12.2%), **Fig. 2e, left**). For the CV models, while they have access to up to 400 available action labels^38^, most labels were irrelevant to the clinical setting, such as “canoeing or kayaking,” “changing wheel”, and “playing musical instrument”. Therefore, the majority of the identified actions among CV models were only distributed among a few labels. For example, MMAction^32^ identified “stand,” “sit” and “lie/sleep” (median (IQR) BD: 55.56% (40.00%), 17.11% (20.89%), 7.11% (7.11%); HC: 42.44% (25.11%), 17.33% (13.55%), 11.33% (13.99%)). Most concerning was that the top three actions detected by S3D^39^ were erroneously identified as “biking through snow,” “folding napkins,” and “folding clothes” (median(IQR): BD: 28.81% (22.27%), 17.17% (23.64%), 13.37% (42.74%); HC: 42.74% (37.16%), 24.55% (30.71%), 10.64% (15.06%)). This was likely due to the mismatch in camera angles between our study and the model’s training data.

In contrast, the motifs obtained from our latent-variable model captured a broad array of interpretable behaviors in the clinical context. Clips from the same motif showed visually similar combinations of actions and activities. Interestingly, our motifs spanned multiple time scales, varying from a few seconds to a couple of minutes, indicating diverse scales of complexity in behavioral dynamics and underlying cognitive processes^40^. A complete list of motif actions is provided in **Table 1**. While some motifs represented intuitively simple activities (e.g., *standstill*), the majority of motifs captured higher-order behavioral sequences that reveal previously undefined actions, even behavioral intentions. For example, motif 1 included a collection of clips related to the *stretch of one body part*, such as *upper body bend*, *arm swing,* and *wrist/ankle rotation*. Motif 4 revealed *fidget*, meaning small movements in hands and feet, such as *nose picking*. In addition, motif 9 showed an active exploratory behavior, in which participants *approached objects and then inspected them*, but did not necessarily directly interact with objects as in motif 8 (**Fig. 2d**). Notably, motif 9 is an intentional exploration, i.e. the subject typically had a targeted object or a destination in mind after scanning around the environment, as opposed to the *aimless wander* in motif 6 and the *depart* after exploration in motif 2.

**Table 1.** Motif descriptions and examples.

| Motif | Description | Examples |
| --- | --- | --- |
| motif 0 | *torso rotation* | *turn walking direction,*<br>*lean left and right,*<br>*bend forward* |
| motif 1 | *stretch (one body part)* | *upper body bend,*<br>*wrist/ankle rotation,*<br>*arm swing* |
| motif 2 | *depart (from the previous action)* | *step away from the window,*<br>*walk away from the desk,*<br>*step away from the bulletin board* |
| motif 3 | *arm and hand movement* | *touch clothes,*<br>*pull open drawers,*<br>*reach objects* |
| motif 4 | *static or fidget* | *pick nose,*<br>*remove the candy wrapper,*<br>*detangle and braid hair* |
| motif 5 | *standstill* | *standstill by the bulletin board,*<br>*standstill in the middle of the room,*<br>*standstill by window* |
| motif 6 | *wander and scan (aimlessly)* | *wander towards the bookcase,*<br>*scan across the room,*<br>*look at the cradle swing* |
| motif 7 | *turn from/to* | *turnaround from the window,*<br>*step back and turn,*<br>*turn head left and right* |
| motif 8 | *examine/interact with objects* | *look at the desk,*<br>*reach objects on the bookcase,*<br>*wear clothes placed on the bookcase* |
| motif 9 | *approach (with aim) and/or inspect* | *approach the bookcase and inspect it,*<br>*go to the door and peek,*<br>*read from the bulletin board* |

To address concerns regarding the distinctiveness of motifs, we conducted an additional analysis comparing key point trajectories across motifs. To evaluate the differences between these variables, we performed a permutation-based two-sample t-test and applied the Benjamini-Hochberg (BH) false discovery rate correction to adjust for multiple comparisons, a procedure applied consistently throughout all analyses in this study. Key point analysis demonstrated that seemingly similar actions, such as “stepping away from the bulletin board” in motif 2 and “turning around from the window” in motif 7, differ significantly in upper body movement dynamics (two-sample t-test BH adjusted p-value: left shoulder-neck, right shoulder-neck, neck-left eye, neck-right eye all < 1 × 10^−10^). Similarly, behaviors like “removing a candy wrapper” in motif 4 differed from general “arm and hand movements” in motif 3 by exhibiting significantly smaller kinematic differences in hand-to-elbow, elbow-to-shoulder, and other key-point pairs (two-sample t-test BH adjusted p-value: left hand-left elbow, right hand-right elbow, left elbow-left shoulder, right elbow-right shoulder, neck-left eye, neck-right eye all < 1 × 10^−10^), justifying why the former is categorized as fidgeting while the latter is not. These quantitative distinctions (**Supplementary Fig. 2**) further validate the model’s ability to disentangle behaviorally meaningful motifs.

The timing and duration of motif occurrences were similar to those of manually annotated labels. For example, as we divided the video into three 5-minute epochs, both approaches showed many behavioral occurrences in epoch 1, and in contrast few occurrences in epoch 3 (**Fig. 2c)**. Although there is not a one-to-one correspondence between manually annotated labels and learned motifs, 87% of the onset and offset of motifs align with those of manually annotated labels (**Methods**). Moreover, motifs displayed a more fine-grained and broader distribution of behavior compared with manually annotated labels. For example, for periods where there is only one human annotated label like “stand,” the latent-variable model has revealed more fine-grained motifs such as *tucking shirts using hands while standing*. This demonstrates that the latent-variable model not only captured the actions that are explicitly perceivable by the eye but also identified finer categories of actions that are data-dependent.

### Differences in Motif Dwell Times Between BD and HC

Our motifs produced relevant representations of human pose for understanding the behavioral characteristics of the euthymic state of BD. People with BD are considered in a euthymic state when they do not meet the criteria for a manic, hypomanic, or depressed episode although they may still exhibit some symptoms. We were interested in whether we could identify distinct behavioral features of euthymic BD patients that distinguished them from HCs, even in the absence of a depressive or manic episode.

To this end, we measured the average motif usage (dwell time) which is the time spent in each motif, for BD and HC during the entire recording period (**Fig. 2e**). Previous work on the hBPM has shown that manic BD patients displayed high motor activity in the first epoch, but quickly attenuated in the second and third epochs^11^. Consistent with this setting, we also calculated the mean dwell time of each *motif* in the three 5-minute epochs.

We detected differences between BD and HC in overall dwell time for motif 1 (*stretch of one body part*), motif 4 (*static or fidget*), and motif 9 (*approach objects then inspect them*) (two-sample t-test p-value; BH adjusted p-value: 0.013, 0.002, 0.022; 0.070, 0.020, 0.073, **Fig. 2e, right**). For clusters obtained by k-means clustering of the key point trajectories, cluster 4 and cluster 6 displayed differences between the populations (two-sample t-test BH adjusted p-value: 0.01, 0.01). In contrast, for manually annotated and CV-identified actions, dwell times associated with their labels did not distinguish between the populations. The dwell time of motifs varied between epochs. We found the dwell time of motif 1 was higher in the BD population in the first epoch (two-sample t-test p-value; BH adjusted p-value: 0.007; 0.080), higher in motif 4 in the third epoch (two-sample t-test p-value; BH adjusted p-value: 0.020, 0.200), lower in BD in motif 9 in the second epoch (**Fig. 2f**, two-sample t-test p-value; BH adjusted p-value: 0.042; 0.173). In the manually annotated categories, “walk” was lower in BD in the last epoch (two-sample t-test BH adjusted p-value: 0.02, **Supplementary Table 2**).

To compare the descriptive power on the distribution of behaviors, we introduced a measure of motif entropy. Specifically, the entropy of the dwell time distributions is the highest for our method (**Supplementary Fig. 3c**). Lower entropy dwell time distributions suggest a model mismatch, as they indicate that most of the probability mass is allocated to a small number of motifs. An ideal fit, according to the principle of maximum entropy, should have a uniform dwell time distribution.

Overall, we found BD patients had more time stretching, fidgeting, and less time in interaction with objects, indicating potential perseveration and impairment of attention^41,42^. However, the differences in dwell time between BD and HC participants were relatively small, suggesting that simply measuring the time a participant spent in each motif is not sufficient to capture complex behavioral dynamics that distinguish between the two populations.

### Motif transitions displayed more stereotypy in BD

Next, we developed more nuanced measures to quantify behavioral dynamics more accurately. Specifically, we focused on two key aspects: (1) the transition frequency between motifs, and (2) the diversity of the behavioral repertoire, as the participants spent more time in the environment. Visual inspection of ethograms highlighted periods during which participants frequently transitioned between motifs, indicating a richer and more diverse behavioral repertoire, in contrast to periods where participants remained consistently within a single motif, or a small subset of motifs (**Fig. 2c**).

To systematically quantify these fluctuations, we viewed motifs as states within a Markov Chain and quantified the temporal relationships between them. We computed the weighted adjacency matrices (*A*) to measure transition frequency and transition probability matrices (*P*) to assess behavioral diversity for each participant (**Fig. 3a, b, Supplementary Fig. 4a-d**). Specifically, adjacency matrices *A* counted how often every motif *S_i_* transitioned to every other motif *S_j_* (where *j* ≠ *i*), and the overall transition frequency was calculated as the sum of all entries in *A* divided by the total number of entries in *A*. The transition probability *P_ij_*, derived from *A*, quantified the probability of each motif *S_i_* transitioning into another motif *S_j_*. These measurements of *A* and *P* were computed at each epoch, allowing us to quantify how frequently individuals shift between different motifs and the likelihood of such transitions occurring. To test the robustness of our results, we computed *A* and *P* with different numbers of motifs (*n* = 10 or 30) to explore the impact of the number of motifs on transition dynamics.

**Fig. 3.**
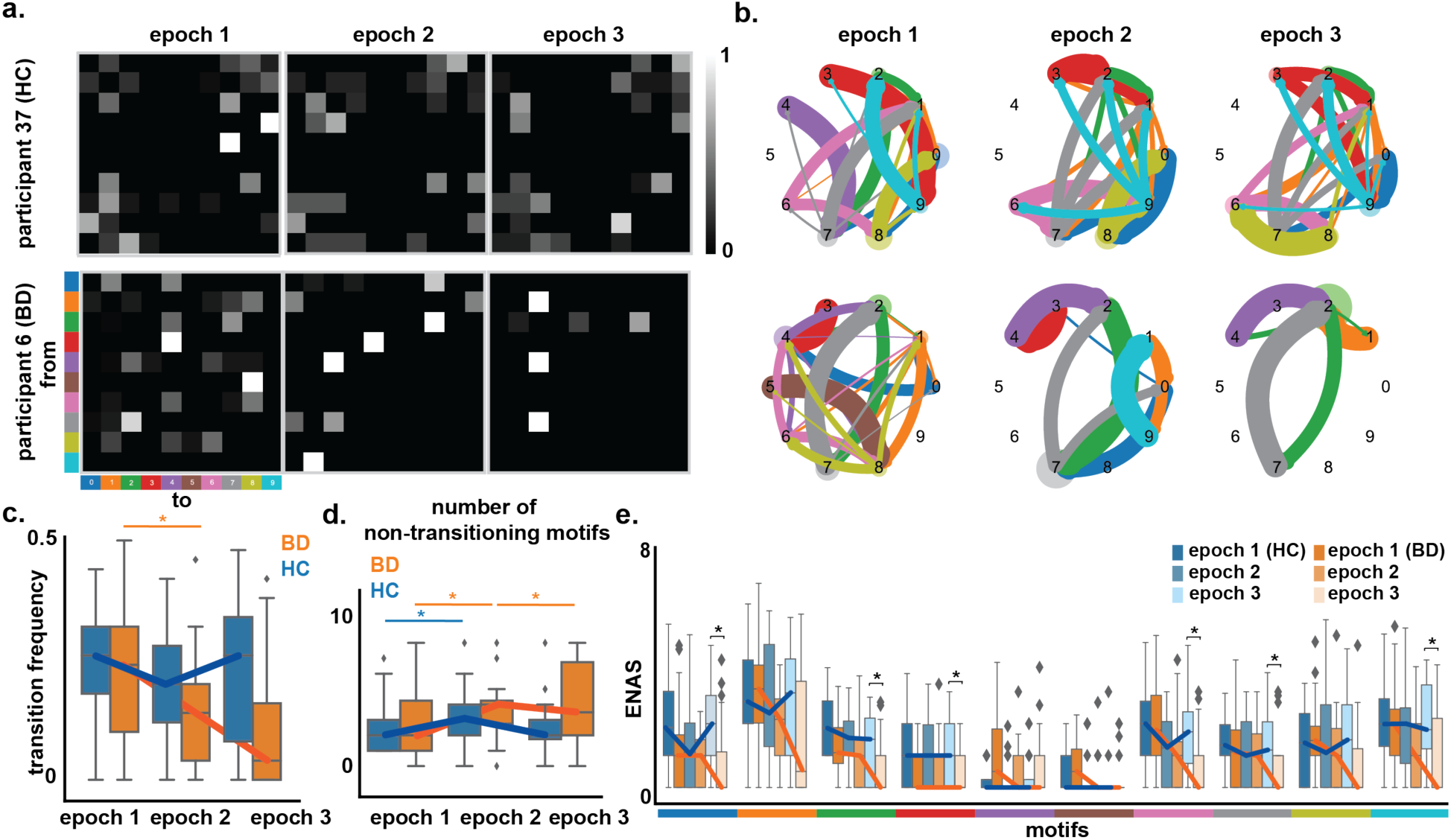
Motif Transition. **a.** Transition matrices in three epochs for an individual HC participant 37 and a BD participant 6, where each pixel represents the transition probability from every motif into every other motif. **b.** Graphs representing the transition matrices in a. where nodes represent motifs and directed edges are colored by the ‘from’ motif color. The thicker the edges the higher the transition probability. The larger the nodes the higher dwell time of the motif. **c.** Transition frequency of three epochs in HC (blue) and BD (orange) with lines connecting the medians in each group over three epochs. **d.** Number of non-transitioning motifs of the HC (blue) and BD (orange) population over time. **e.** Effective-number-of-accessible-states (ENAS) of three epochs of HC (blue) and BD (orange) of ten motifs. Epoch 1 – epoch 3 marked by dark to light shades in each population. Significance marked by asterisks.

While both BD and HC groups experience an overall decrease in transition frequency, the decline is more pronounced in BD patients between epoch 1 and 2 (**Fig. 3c**, paired t-test p-values epoch 1 vs epoch 2, epoch 2 vs epoch 3, BD: 0.002, 0.118, HC: 0.086, 0.408). We further quantified sparsity of the transition matrices by counting the number of non-transitioning motifs, in which a participant remained “stuck” without ever transitioning to other motifs, by counting the rows in the transition matrices that contained only zeros, while the corresponding dwell time is nonzero (**Fig. 3d**). This measure reflects whether behavior was dominated by a set of isolated, stereotypical states with no observed transitions. We found a significant increasing trend in non-transitioning motifs in BD between all epochs, but only between epoch 1 and 2 in HC (paired t-test p-value epoch 1 vs epoch 2, epoch 2 vs epoch 3, BD: 0.002, 0.025, HC: 0.005, 0.813)

While the number of non-transitioning motifs captures extreme cases of behavioral rigidity, it does not account for the overall structure of transitions between motifs. To further quantify stereotypy beyond these states, we introduced a novel measure, the *effective-number-of-accessible-states* (*ENAS*) of the transition matrix. *ENAS* is a measure of the number of accessible motifs (states) for each period (overall time, or epoch) by weighting the count of motifs by their relative accessibility (probability). Intuitively, *ENAS* reflects how freely a participant moves between different behavioral motifs. If a participant transitions equally to all other motifs from a given motif *i*, the *ENAS* for that motif equals to maximum value (*n* − 1, where *n* is the total number of motifs). Conversely, if the participant remains “stuck” in the same motif without transitioning to others, the *ENAS* is 0, indicating highly stereotyped behavior; if a motif deterministically transitions to only one other motif, then ENAS is 1 (See **Methods** for full details).

We found *ENAS* became smaller for BD patients over time in all motifs; within the same epoch, *ENAS* often was smaller for BD patients compared with HC participants, especially in epoch 3 (motifs 0, 2, 3, 6, 7, 9 two sample t-test BH adjusted p-value: 0.03, 0.04, 0.04, 0.03, 0.04, 0.04, **Fig. 3e**). This indicated a decrease in behavioral repertoire in BD behavior over time. *ENAS* provided a way to quantify the decline in *activation* in BD, a multifaced construct often associated with many overlapping constructs such as *psychomotor retardation* and *disengagement*^43^. Importantly, a decline in *activation* did not imply *inactivity* (i.e., no change in key point positions). The decreasing *ENAS* suggested that while BD individuals continued to engage in behaviors, their behavioral repertoire became progressively constrained to a subset of accessible motifs, suggesting diminished activation and increased stereotypy over time.

We found that this pattern of increasing stereotypy over time remained true even when using a three-fold increase in the number of motifs (*n* = 30, **Supplementary Fig. 4 e-f**), The fact that these results are robust to the exact model specification indicates that the observed increase in stereotypy over time is a hallmark of BD, and not idiosyncratic to our model. Moreover, our analysis of transitions highlights the need for more sophisticated quantification methods such as *ENAS*, as subtle changes in behavioral flexibility and stereotypy may not be fully captured by simply assessing the duration spent in each behavior. Altogether, our results suggest that BD patients’ behavior tends to become more stereotyped, and decrease in *activation* during the course of recording, even during euthymic episodes.

### Latent representations displayed behavioral variability in BD

Transition analysis captured the dynamics between motifs; however, it did not account for the diversity of actions that can occur within the same motif. For example, in motif 1, one participant may stretch by *rolling their arms*, while another may *kick their legs*. To quantify this within-motif variability, we used a measure of *motif-volume*. Intuitively, actions that were physically similar were represented by trajectories that lie close together in the latent space. Therefore, the spread of latent variables reflected the variability of movement within a motif. We define *motif-volume v*_*i*_(*τ*) as the total variance of the latent representation of motif *i* at time *τ*. A larger *motif-volume* indicates greater variability in how a motif was expressed across participants, while a smaller *motif-volume* suggests participants exhibit more uniform behaviors within that motif.

We observed BD *motif-volume* was consistently decreasing over time in more than half of the motifs (**Supplementary Fig. 5a, b**) and became significantly lower than HC in epoch 3 in motif 0 (*torso rotation*), 2 (*depart*), 6 (*wander and scan*) and motif 9 (*approach objects then inspect them*, two sample t-test BH adjusted p-value of between BD and HC epoch 1-3, motif 0: 0.03, 0.03, 0.03; motif 2: 0.72, 0.21, 0.04, motif 6: 0.83, 0.83, 0.04, motif 9: 0.83, 0.33, 0.03, **Fig. 4b**). This suggested that while BD and HC populations demonstrated a marginal-to-no statistical difference in dwell time, the variability in the expression of this motif tended to be smaller in BD patients. *Motif-volume* in BD was consistently higher than HC in motifs 4 (*static or fidget*) and 5 (*standstill*, two sample t-test BH adjusted p-value of epoch 1-3, motif 4: 0.006, 0.276, 0.078, motif 5: 0.191, 0.935, 0.485, **Fig. 4a, b**), suggesting a larger, and more varied fidgeting and static behavior expression in BD patients. Notably, *motif-volume* is not necessarily correlated with dwell time (**Supplementary Table 4**), indicating that volume is not merely a consequence of more time spent in a given motif.

**Fig. 4.**
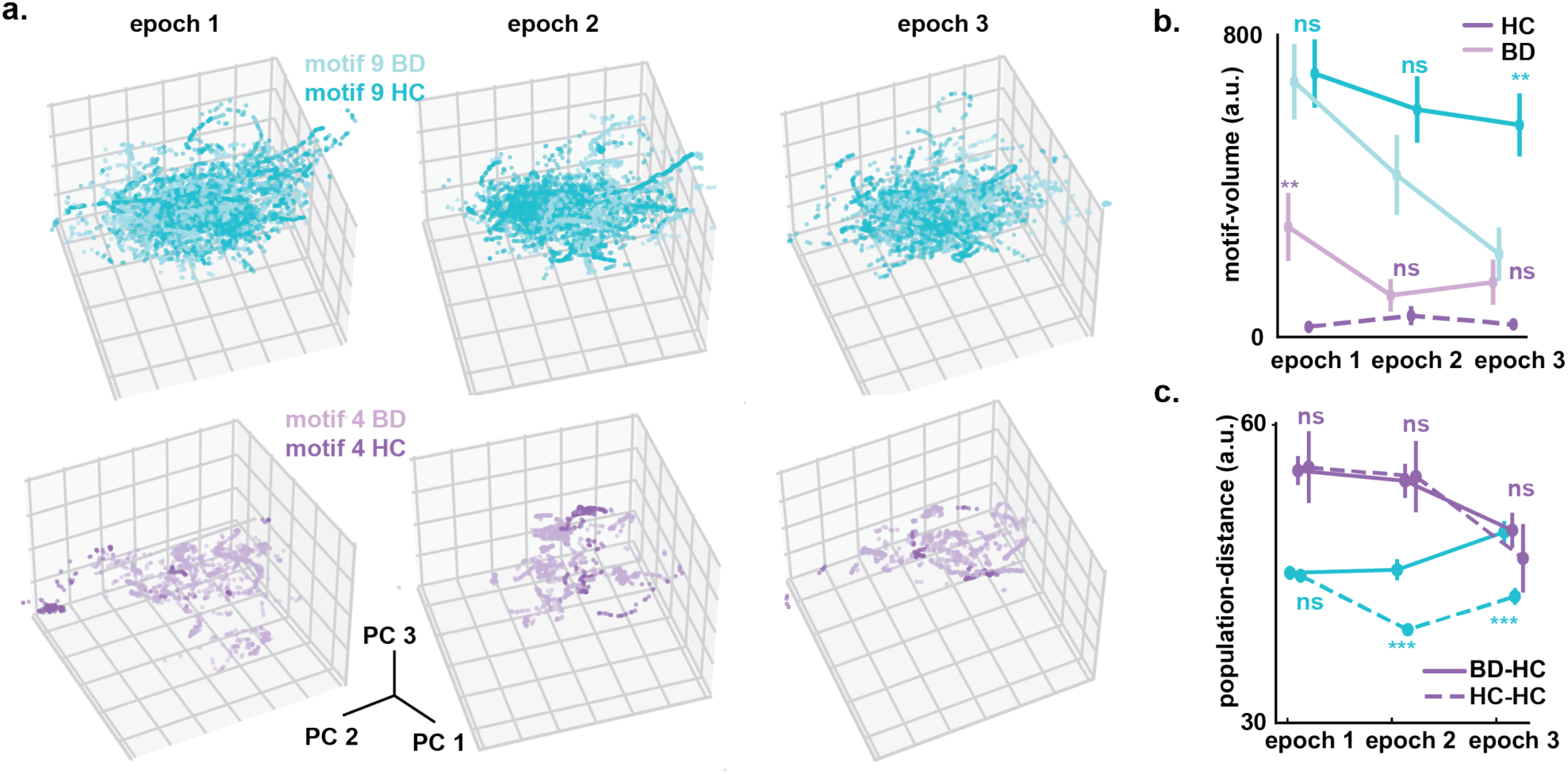
Latent Shifting of Motif Representation. **a.** motif 9 and motif 4 of BD (lighter shades) and HC (darker shades) latent vector in three epochs represented in the top three PC. Latent vectors were shuffled in index and subsampled for visualization. **b.** *Motif-volume* over time for motif 9 and motif 4 in BD (lighter shades) and HC (darker shades) population. **c.** Interpopulation-distance between BD and HC (solid lines) in epoch 1, epoch 2, and epoch 3. As control, intrapopulation-distance of HC (dashed lines) were shown. Significance was marked by asterisks.

To quantify within-motif variability between populations over time, we computed the interpopulation distance between BD and HC latent representations of each motif in each epoch. As a control, we computed the intrapopulation distance within BD, and within HC in each epoch (**Fig. 4c**). If latent representations are dissimilar between BD and HC, then the interpopulation distance would be larger than expected by chance and the volumes representing the motif for both populations would overlap less.

We found while the interpopulation distances changed over time across different motifs (**Supplementary Fig. 5c**), the interpopulation distance was larger than the HC-intrapopulation distance in many motifs, especially in the last epoch (two sample t-test motifs 1, 2, 5, 6, 7 and 9 BH adjusted p-values all < **0. 011**). This indicated BD and HC individuals expressed these motifs with increasingly distinct patterns of movements. Together, motif-volume and interpopulation distance quantitatively assessed the divergence of behavior expression within individual, capturing subtle behavioral differences that temporal features did not reveal. These findings not only highlighted the progressive divergence between BD and HC but also suggested that BD may be associated with the development of more stereotypical and more distinct behavior, providing a potential avenue for monitoring disease symptoms.

### Behavioral features from the latent space better discriminate BD from HC than traditional measurements

The behavioral features we derived from the segmented latent representations of actions are consistent with the phenotype of increased activity and energy, which is a hallmark feature of BD. These features arguably encompassed a less biased set of behavioral markers of BD compared to CV models, expert human annotation, and even established clinical assessment scales as they were discovered from spontaneous human behavior in real-world contexts, rather than pre-defined catalogs of behaviors. We thus hypothesized that the identified behavioral features would better distinguish euthymic BD participants from HCs, than alternative methods.

To evaluate the ability of our behavioral features to distinguish euthymic BD participants from HC participants, we trained a logistic regression classifier using features selected from both assessment scales (HAM-D, YMRS) and behavioral features. Feature selection was performed using 4-fold cross-validation within the training set to ensure generalizability. We found the most predictive features of BD were *motif-volume* and *ENAS* (**Supplementary Table 3**). To assess classification performance in a subject-independent manner, we employed leave-N-subject-out cross validation (N=5), where N participants were held out as the test set while the model was trained on the remaining participants. As benchmarks, we compared classification performance against models trained on (1) psychometric assessment scales, (2) human-annotated behavioral labels, and (3) features from CV-based models (**Table 2**). We found that the classification accuracy using our behavioral features outperformed all of the other approaches (Tukey HSD p-value Ours vs other approaches in Table 2 order all < 1 × 10^−3^). Our results underscore the potential of data-driven behavioral motifs to differentiate BD from HC effectively.

**Table 2.** Classification accuracy of BD vs HC across approaches.

| model | pretrained | accuracy (%) |
| --- | --- | --- |
| Assessment Scales | - | 37.41 |
| Spatial-D | - | 33.33 |
| K-means on DLC | - | 71.48 |
| hBPM video ratings | - | 71.11 |
| S3D | Kinetics-400 | 63.70 |
| MMAction2 | Kinetics-400 | 72.96 |
| **Ours** | - | **77.04\*** |

Finally, to investigate whether the differences observed in BD vs. HC populations are influenced by medications, we performed independent t-tests on the features discussed above for three comparisons: BD (all) vs. HC, BD (no medications, n=8) vs. HC and BD (no medications) vs. BD (on medications) and applied BH procedure to correct for multiple comparisons. The results (**Supplementary Table 5**) reveal that 14 features exhibit significant differences between BD (all) and HC, particularly in *motif-volume*, *ENAS*, and dwell time features, suggesting robust group-level effects. While BD patients without medications showed significant differences in 8 *motif-volume* and *ENAS* features compared to HC, patients on medications were not significantly different from patients without medications across 13 features. Thus, although BD (no medications) exhibited significant differences from HC in only a subset of the features distinguishing BD and HC, this may merely reflect a considerably smaller sample size of the no-med BD sample (8-no meds vs 24-all), rather than the impact of medications per se. Indeed, taken together we regard these results to reflect robust and multifaceted differences across BD and HC groups indicating that many features derived from our method independently enhance the discriminability of BD from HC subjects.

To account for potential confounding effects of age, we ran ANCOVA (Analysis of Covariance) for each feature using an Ordinary Least Square (OLS) regressing with age as a covariate. Of the 35 features originally distinguishing BD from HC, 12 remained significant (p < 0.05), suggesting that these differences were not explained by age. An additional 2 features showed marginal significance (p < 0.06), indicating a potential trend that may be influenced by sample size or individual variability. As mentioned above, one limitation of our study is the sample size (n=48), which while relatively large for a translational human study, may limit the generalizability. Another consideration is that BD participants were on various medications, potentially influencing behavioral measures. Future studies should further examine pharmacological influences through controlled longitudinal designs.

## Discussion

Current data-driven machine learning techniques may offer significant advantages over traditional observational methods across a wide range of domains, as the latter methods are potentially prone to bias. While recent advances in computational methods have laid the groundwork for extracting stereotyped motifs from continuous animal behavior, their application to understanding more complex human behaviors in psychiatric populations remains underexplored. By integrating computer vision, deep learning, and probabilistic reasoning to study activation in BD, we present a novel approach for identifying subtle behavioral patterns in clinical settings. Our findings provide a foundation for applying unsupervised machine learning to psychiatric behavior analysis, demonstrating that data-driven approaches can uncover meaningful behavioral signatures that elude traditional assessments and overcome the limitations of human annotation. Our framework offers a promising avenue for assisting diagnostic precision, informing treatment strategies, and bridging the gap between human and animal models of neuropsychiatric disorders.

Through an end-to-end design, we identified behavioral motifs and transition dynamics that differentiate euthymic BD individuals from healthy comparisons— a task that is particularly challenging due to the complexity and subtlety of behavioral differences in BD. We validated our model using a classification task to distinguish euthymic BD from healthy comparisons, based on latent representations and features of the motifs (e.g., transitions between behaviors, stereotypy), benchmarked against rating scales, traditional action recognition and manual annotations. Our model outperformed all of these alternatives, indicating that it captured clinically relevant behavioral information in BD. More broadly, our findings suggest that a more precise characterization of the clinical presentation of the participants has been learned by the model and can be used in various downstream tasks that could offer valuable insights for clinical assessment and treatment planning.

Our unsupervised method minimizes human bias in behavioral categorization, enabling the discovery of clinically meaningful motifs by capturing fine-grained subtle patterns that may not fit conventional diagnostic scales. This improvement is particularly relevant in psychiatry, where symptom presentation varies across individuals and traditional assessments may fail to capture fine-grained behavioral signatures. For example, our analysis revealed that BD participants had reduced dwell times when *approaching some objects and inspecting them*, suggesting differences in attentional engagement^41^. This aligns with previous studies showing impaired working memory and attention in euthymic BD^44,45^. However, the direct relationship between these behavioral differences and cognitive function in BD remains to be explored. Future research should examine whether these behavioral patterns are linked to broader cognitive functions or impulsivity-related deficits in BD^46^. Additionally, more fidgeting movements, such as *tapping feet* or *scratching hair*, were observed in BD patients in our study. While the clinical significance of these behaviors requires further investigation, prior studies have linked similar patterns to perseverative behavior in manic and hypomanic BD patients^36,42^. Notably, these behaviors were not listed in established behavior rating criteria or detected by both general-purpose action detection software and human annotators.

However, while our motifs offer more nuanced behavioral insights, their clinical utility remains a challenge. Linking motifs to symptoms manifested in mania and depression in BD— for example by tracking motif expression alongside symptom fluctuations—is an interesting future direction for further study. For example, integrating our approach with a passive, real-world monitoring wearable device could enable continuous behavioral assessment beyond controlled experimental settings. For example, exploring whether specific motifs can predict real-world functional outcomes, such as sleep quality or the ability to perform tasks of daily living, could strengthen the clinical value of our approach and deepen our understanding of functional impairments in BD. Additionally, our controlled experimental real-world setting ensured consistency but may not fully capture real-world behavior complexity. Expanding our framework to naturalistic environments will enhance its translational relevance.

The motif-based analysis can enable cross-species comparisons, offering insights into shared neurobehavioral mechanisms and deepen our understanding of psychiatric symptomatology. For instance, human *fidgeting* may be analogous to rodent *grooming* behaviors, reflecting common responses to environmental stressors or internal states. This cross-species perspective is particularly relevant in translational research, where behavioral phenotyping in animal models is critical for understanding psychiatric conditions and testing therapeutic interventions. Furthermore, while the focus of our study was on BD, future research may explore whether this approach can aid in differential BD versus unipolar depression. More broadly, our approach is diagnosis-agnostic and modular, making it adaptable to other neuropsychiatric conditions.

## Data Availability

All data produced in the present study are available upon reasonable request to the authors

## Acknowledgements

We thank Kristen Kraffert, Haili Song, and Nathan Wood for their help in data collection.

Zhanqi Zhang is supported by the HDSI Ph.D. Fellowship at the University of California San Diego. Chi K. Chou was supported by the Shenoy Research Mentor Fellowship in Neuroscience (SURFiN).

This study was conducted under IRB #180344 and was supported by NIH grants NIDA R01DA043535 (W.P./J.Y.) and NIDA R01DA051295 (A.M./J.Y.).

## Contributions

A.M., J.Y., and W.P. designed the experiments and collected the data. Z.Z, G.M., and M.A. conceptualized the experiment analysis and analyzed the data with assistance from C.C., and H.R. Z.Z wrote the manuscript under G.M. and M.A.’s supervision. Z.Z., G.M., M.A., A.M., J.Y., and W.P. reviewed and edited the manuscript.

## Methods

### Data and Procedure

All Patients (n = 24; 10 men) were between the ages of 19 to 56. Among the population, all but one patient was diagnosed with bipolar disorder (BD) Type I or Type II(defined by the Structured Clinical Interview for DSM-IV^37^). The remaining patient was diagnosed with the cyclothymic subtype of BD. All BD participants were in a current euthymic episode. Non-patient participants (n = 24; 12 men) of matching years of age who had never met the DSM-IV^37^ standard for alcohol or substance abuse or dependence, tested positive on a urine toxicology screen, had a neurological ailment, or had a condition affecting their motor skills were recruited for the study as the healthy control group (HC). Participants from both BD and HC populations were evaluated with the Young Mania Rating Scale (YMRS)^4^ and Hamilton Depression Rating Scale (HAM-D)^2^, and all BD and HC participants had YMRS < 12 and HAM-D < 10. Most of the BD patients were treated with one or a combination of mood-stabilizing, antipsychotic, antidepressant, and sleep aid medication; other BD patients were not on medication during testing. See **Supplementary Table 1** for full information.

Participants consented to have their activities filmed during an unspecified segment of the research session. The video data was collected at the UCSD Medical Center in an unused office room that was designed to appear in transition. The room was 2.7 m × 4.3 m with a periphery lined with various pieces of furniture, such as a desk, both small and large filing cabinets, and two sets of bookshelves. No furniture that could directly lead to sedentary behavior was set in the room. Eleven small objects were placed evenly on items of furniture. These items were selected based on the condition that they are safe, vibrant, tactile, easily handled, and are likely to encourage exploration by humans^50^.

Participants were directed to wait in the room with minimal instructions until the examiner returned. Participants were not allowed to leave the room or bring personal items into the room. The videos were recorded for *T* = 15 minutes continuously from a commercial camera with a fisheye lens hiddenly placed at the center of the ceiling. The recordings had a resolution of 640 x 480 pixels and a frame rate of 30 frames per second. Following the procedure in the previous studies on the dataset^10,11^, the recorded session of 15 minutes was evenly divided into three 5-minute epochs for analysis in this study.

Human experts reviewed the video recordings afterward to count instances of 11 exploration action categories, including sitting with or without an object, standing with or without an object, walking with or without an object, lying with or without an object, wearing an object, exercising, and interacting with objects such as drawers and window blinds^11^.

The spatial scaling exponent (Spatial-d) estimated the geometric structure of the path of the participants, first introduced in animal behavior studies^51^ and used as a metric in previous human behavior studies on this dataset. It estimates the linear slope of *log*(*L_k_*) with respect to *log*(*k*) where *L_k_* is the average length of the path and *k* is the measuring resolution of the movements.

### Human Pose Tracking and Estimation

Existing methodologies for human motion tracking were not developed for a single top-view camera with fish-eye distortion and thus performed poorly on this dataset. To characterize the participant’s behavior, we used DeepLabCut^15^. Specifically, in DeepLabCut we first clustered the frames using k-means and selected frames from different clusters to obtain 20 - 50 frames from each video. This process ensures that the selected frames cover different poses of the person. We labeled these frames with markers at 20 anatomical landmarks (left eye, right eye, left ear, right ear, mouth, the center of the neck, left shoulder, right shoulder, left elbow, right elbow, left hand, right hand, the center of hip, left hip, right hip, left knee, right knee, left foot, right foot, the center of feet). The labeled frames were used for training a ResNet-50^52^ model to learn and predict marker position in the remaining frames. In order to have accurate marker estimation, the training involved 3 iterations, with 1,030,000 epochs each. After each iteration, 10 outlier frames (DeepLabCut confidence score below 0.1) with inaccurate marker estimates from every video were relabeled and added to the training set for the next iteration. Training iterations were terminated when the training and testing errors of the DeepLabCut marker estimation were 2.03 pixels and 3.71 pixels, respectively. The x-y position estimates of the 20 body parts for each frame were used for subsequent analyses.

### Key Point Marker Postprocessing

Key point markers of all subjects were aligned to egocentric coordinates. To accomplish this, we cropped the frame to fit the subject within a bounding box (300 x 300 pixels). Then we aligned the skeleton using the key points of the center of the hip, and center of the feet markers as reference. As a result, the upper body markers were located at the top of the cropped frame, and the lower body markers at the bottom. Marker estimates with less than 90% confidence level determined by DeepLabCut were removed.

### Encoding the Pose into Latent Space

To identify distinct behavioral motifs from times series of pose coordinates, we adapted the pipeline in the Variational Animal Motion Embedding (VAME) model^22^, which has been used previously to identify open-field mouse behaviors using a bidirectional RNN variational autoencoder (VAE) and clustering. The VAME model was used to encode and reduce the dimensionality of the pose sequence of the human participants. Specifically, the latent dimensionality was set to *d* = 10, a value less than the input dimension of 40 (20 markers with x and y coordinates). The resulting latent representation *Z* for each subject is thus a matrix of size *d* × *T*.

The original VAME model used a hidden Markov model for extracting 50 motifs of the animal, used hierarchical clustering of motifs to obtain a tree-structured graph, and then grouped motifs into communities by cutting the tree at a certain level/depth of the branches. However, because human behavior may be more complex, the hierarchical representation of human behavior varied across motifs and was not visually similar in each community. We instead performed k-means clustering on the latent representation to obtain the behavioral motifs. As a direct comparison with 10 labels from human annotation, we included the results of 10 clusters in the main results of this study. We also reproduced our analysis using k = 30 clusters with results included in **Supplementary Fig. 4e, f**.

### Matching Annotation Labels with Motif Labels

For each video, we obtained a list of human annotations and a list of motif labels. Since the labels from human annotations and motifs obtained from the latent-variable model do not necessarily match one-to-one, we measured how many times the onset and offset of each label matched between the two labels. Both lists were filled with integers representing the action labels at each frame. For example, the first 8 frames from one video may be represented as `[a, a, a, b, b, b, c, c]`, with `a`s`, b`s`, c`s standing for the labels of the action on that frame. We first divided the lists into chunks `[a, a, a], [b, b, b], [c, c]` so that each chunk represented an epoch with only one label, and a delimiter ‘0’ was added between chunks. The output of the example frames would be `[[a, a, a], 0, [b, b, b], 0, [c, c]].` Since the objective was to find the onset/offset alignment, which was marked by the location of the `0`s only, the labels could be simplified as `[[1, 1, 1], 0, [1, 1], 0, [1, 1, 1]], with 1s` representing the chunks of labeled frames while `0` representing the chunk boundaries.

We computed the total number of chunks in human annotations, and the number of matching chunks between human annotation and motif labels in terms of onset/offset timestamp. Because human annotations of onset and offset of actions had inherent uncertainty, we defined a specified `offset` value allowing for a certain number of frames of mismatch.

For example, between

`list1 = [0, 1, 1, 1, 1, 1, 1, 0, 1, 1]` and

`list2 = [1, 0, 1, 1, 1, 0, 1, 1, 1, 0],`

with an offset of `2`, there are two matching labels chunks: `[1, 1, 1, 1, 1, 1]` with `[1, 1, 1]` and `[1, 1]` with `[1, 1, 1]`. We reported the ratio of matching labels to total human annotation labels. There are 34.09% of labels that were matched when the offset was 1 second, 77.54% when the offset was 5 seconds, and 87.60% when the offset was 10 seconds.

### Computing effective-number-of-accessible-states (ENAS)

Each *i* ∈ *n* row of the transition matrix *P* is composed of the transition probability, *P_ij_* from motif *S*_*i*_ into every other motif *S*_j_. The intuition behind the *ENAS* is to measure how many motifs could be accessible based on the current observed transition matrix. If 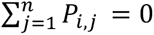, this indicates no other motif was visited from motif *i*, resulting in *ENAS* of motif *i* to be 0 (self-transitions were excluded from computations). Otherwise, we compute *ENAS* of the motif *i* in the following manner.

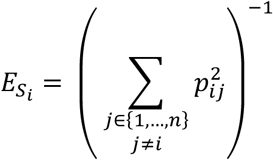

The 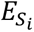 represents the number of accessible motifs from the current motif *i*, which is a number between 0 to *n* − 1, where *n* is the number of total motifs. If there is no motif accessible from the current motif, then 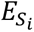 will be 0.

The overall ENAS *E* is the average of 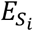 over all motifs *S_i_* for *i* ∈ {1, …, *n*}

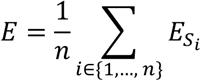

The pseudo-code for ENAS is the following:

<u>ENAS(P):</u>

for *row_i_* in *P*:

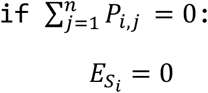

else:

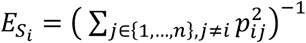

### Computing Volume and Distance of Latent Representations

To compute the *latent-volume*, we first mean-centered the latent vectors of all motifs during the entire time *T*. The *latent-volume v*_*i*_(*τ_m_, p*) of the latent representation 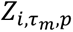 of *motif i* at the time *τ_m_* of population, *p* was quantified by the trace of the covariance of the latent vector *Z*_*i*_

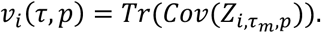

To compute the *population-distance*, let’s define the following:

At each motif *i* ∈ [1, 2, …, *k*] and during each epoch τ*_m_*, the latent representation of a BD subject to be *X_i_*, *τ_m_* of ℝ*^d^*, and the latent representation of an HC subject to be 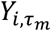 of ℝ*^d^*, where *d* is the latent dimension.

Assume 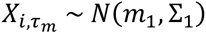 and 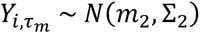, meaning each point in 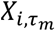 and 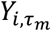 is an independent sample from its respective Gaussian distribution, with expected values and covariance.

We computed the 2-Wasserstein distance between 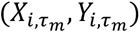 at each motif *i* ∈ [1, 2, …, *k*] and during each epoch *τ_m_*. Specifically,

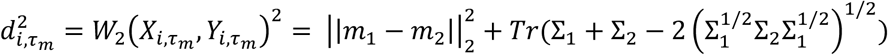

where, *m*_1_*, m*_2_ and Σ_1_, Σ_2_ are sampled means and covariances. The 2-Wasserstein distance was computed with the Python function below.

*Interpopulation-distance* was the mean of pairwise 2-Wasserstein distance between every subject in BD and every subject in HC. For comparison, we computed *intrapopulation-distance*, as the mean pairwise 2-Wasserstein distance within the HC group and within the BD group.

~~~
def wasserstein_distance(m1, C1, m2, C2):
     ““”
     Calculate the 2-Wasserstein distance between two Gaussian distributions.
     Parameters:
     m1, m2: Mean vectors of the two Gaussian distributions (numpy arrays).
     C1, C2: Covariance matrices of the two Gaussian distributions (numpy arrays).
     Returns:
     W2: The 2-Wasserstein distance.
     ““”
     # Euclidean distance between the means
     mean_diff = np.linalg.norm(m1 - m2)
     # Principal square roots of the covariance matrices
     # Calculate the trace term
     term = sqrtm(sqrtm(C2) @ C1 @ sqrtm(C2))
     trace_term = np.trace(C1 + C2 - 2 * term)
     # Wasserstein distance squared
     W2_squared = mean_diff ** 2 + trace_term
     return np.sqrt(W2_squared).real
~~~

### Visualization of the Latent Representation

Since the latent representation is in a dimension of *d* × *T*, we transformed the latent space using PCA, and the first three principal components (PCs) were plotted for visualization purposes. The motif centroids and centroid distances defined above were also computed separately in PC space and plotted in the top three PCs for proper visualization. All latent representations were visualized in the PC space (computed from the entire latent representation).

### Baseline Computer Vision Models

We selected two state-of-the-art computer vision action recognition models, MMAction2^32^ and S3D-CNN^39^ since not many models would detect the person in the setting of the top view fisheye camera used in the study.

We adapted OpenMMLab’s official repository for MMAction2 (https://github.com/open-mmlab/mmaction2). MMAction2 consists of two modules: a human detection using faster RCNN ResNet50 with COCO dataset, and an action detection using SlowFast ResNet50 network pretrained on Kinetics-400 first for action classification and then fine-tuned on AVA v2.2 dataset for person detection. All pretrained weights and configuration files were downloaded from the repository. We used the following configuration and checkpoints for MMAction2:

~~~
    --config
    configs/detection/ava/slowfast_kinetics_pretrained_r50_8x8x1_cosine_10e_ava22_rgb.py
    --checkpoint slowfast_kinetics_pretrained_r50_8x8x1_cosine_10e_ava22_rgb-b987b516.pth
    --det-checkpoint faster_rcnn_r50_fpn_mstrain_3x_coco_20210524_110822-e10bd31c.pth
    --det-score-thr 0
    --action-score-thr 0
    --label-map tools/data/ava/label_map.txt
~~~

For S3D, we used the unofficial PyTorch implementation (https://github.com/kylemin/S3D), which was pretrained on the Kinetics-400 dataset with pretrained weights downloaded from the same repository. S3D takes in the video dataset and outputs the labels from Kinetics-400 for each frame in the video.

### Selecting Features for Classification

Our data is comprised of numerical input features and categorial output labels (BD and HC). We applied backward feature selection using `SequentialFeatureSelector(n_features_to_select=30, direction=“backward”,scoring=’accuracy’, cv=4)` from `sklearn.feature_selection`. This is a greedy sequential feature algorithm that sequentially removes features from all features based on a 4-fold cross-validated score of the accuracy of the logistic regression classifier. The feature selector stops removing features when the desired number of selected features is reached. Before feature selection, there are 94 input features of each human video, including each motif’s dwell time at three epochs, ENAS of each motif at three epochs, ENAS of all motifs at three epochs, number of zeros in transition matrices, motif volume at three epochs, YMRS scale, and HAMD scale. After feature selection, 30 features were selected from each approach (**Supplementary Table 3**).

### Classifying BD from Behavior Features

Selected features were fed into a binary logistic regression classifier. We utilized a logistic regression classifier from scikit-learn (`LogisticRegression`) with a maximum number of iterations set to 1000. Each feature of the dataset was min-max scaled using `MinMaxScaler` from `sklearn.preprocessing`. For each iteration, we split the data randomly into 75% training and 25% testing sets using stratified sampling, then trained a logistic regression classifier for each iteration, and computed accuracy, precision, and recall scores (using the `accuracy_score`, `precision_score`, and `recall_score` functions from scikit-learn) on the test set for each iteration. We conducted leave-N-subject-out-cross validation, where N=5 participants were held out and the classifier was trained on the remaining 43 participants. We reported mean and standard deviation of accuracy, precision, and recall scores across all iterations. We performed Tukey’s HSD (honestly significant difference) test between pairwise scores between our model and other models with multiple comparison consideration and reported the p-values.

## Supplementary

**Supplementary Table 1.** Participant demographic table and medications.

| **Participant**<br>**s** | **Psychotropi**<br>**c**<br>**Medications** | **Group** | **BD**<br>**Episod**<br>**e** | **Age**<br>**rang**<br>**e** | **Gende**<br>**r** | **Race** | **Ethnicit**<br>**y** |
| --- | --- | --- | --- | --- | --- | --- | --- |
| **participant 1** | mood stabilizer;<br>antipsychotic;<br>antidepressant;<br>sleep aid | BD | Euthymic | 34–38 | Female | White | Non-Hispanic |
| **participant 2** | None | BD | Euthymic | 44–48 | Male | White | Non-Hispanic |
| **participant 3** | no psychotropic<br>medicines | BD | Euthymic | 24–28 | Trans<br>Male | White | Hispanic |
| **participant 4** | no psychotropic<br>medicines | BD | Euthymic | 44–48 | Male | White | Hispanic |
| **participant 5** | no psychotropic<br>medicines | BD | Euthymic | 24–28 | Female | White | Non-Hispanic |
| **participant 6** | 2 mood<br>stabilizers; 2<br>sleep aids | BD | Euthymic | 34–38 | Female | White | Non-Hispanic |
| **participant 7** | mood stabilizer;<br>antipsychotic | BD | Euthymic | 34–38 | Male | White | Non-Hispanic |
| **participant 8** | no psychotropic<br>medicines | BD | Euthymic | 29–33 | Male | White | Hispanic |
| **participant 9** | no psychotropic<br>medicines | BD | Euthymic | 44–48 | Female | Black or<br>African<br>American | Non-Hispanic |
| **participant 10** | 2 mood<br>stabilizers; sleep<br>aid | BD | Euthymic | 44–48 | Female | White | Hispanic |
| **participant 11** | mood stabilizer | BD | Euthymic | 19–23 | Male | White | Non-Hispanic |
| **participant 12** | None | BD | Euthymic | 39–43 | Male | White | Non-Hispanic |
| **participant 13** | antipsychotic | BD | Euthymic | 29–33 | Female | Asian | Non-Hispanic |
| **participant 14** | antipsychotic | BD | Euthymic | 44–48 | Female | White | Hispanic |
| **participant 15** | mood stabilizer;<br>antipsychotic;<br>sleep aid | BD | Euthymic | 39–43 | Female | White | Non-Hispanic |
| **participant 16** | mood stabilizer;<br>antipsychotic;<br>antidepressant | BD | Euthymic | 54–58 | Female | White | Non-Hispanic |
| **participant 17** | mood stabilizer;<br>antidepressant | BD | Euthymic | 39–43 | Male | Black or<br>African<br>American | Non-Hispanic |
| **participant 18** | mood stabilizer;<br>antipsychotic | BD | Euthymic | 39–43 | Female | White | Non-Hispanic |
| **participant 19** | Antipsychotic;<br>antidepressant | BD | Euthymic | 34–38 | Female | American<br>Indian/Alaska<br>n Native | Non-Hispanic |
| **participant 20** | antipsychotic | BD | Euthymic | 44–48 | Male | White | Non-Hispanic |
| **participant 21** | 2 mood<br>stabilizers;<br>antipsychotic;<br>antidepressant | BD | Euthymic | 24–28 | Female | White | Non-Hispanic |
| **participant 22** | 2 antipsychotics;<br>2 sleep aids | BD | Euthymic | 39–43 | Female | Black or<br>African<br>American | Non-Hispanic |
| **participant 23** | mood stabilizer | BD | Euthymic | 19–23 | Female | Black or<br>African<br>American | Non-Hispanic |
| **participant 24** | no psychotropic<br>medicines | Cyclothymi<br>c | Euthymic | 34–38 | Male | White | Non-Hispanic |
| **participant 25** | no psychotropic<br>medicines | HC | N/A | 34–38 | Male | White | Non-Hispanic |
| **participant 26** | no psychotropic<br>medicines | HC | N/A | 29–33 | Male | White | Non-Hispanic |
| **participant 27** | no psychotropic<br>medicines | HC | N/A | 24–28 | Female | Black or<br>African<br>American | Hispanic |
| **participant 28** | no psychotropic<br>medicines | HC | N/A | 24–28 | Female | White | Non-Hispanic |
| **participant 29** | no psychotropic<br>medicines | HC | N/A | 29–33 | Female | White | Non-Hispanic |
| **participant 30** | no psychotropic<br>medicines | HC | N/A | 24–28 | Trans<br>Male | White | Hispanic |
| **participant 31** | no psychotropic<br>medicines | HC | N/A | 24–28 | Male | White | Hispanic |
| **participant 32** | no psychotropic<br>medicines | HC | N/A | 24–28 | Male | Black or<br>African<br>American | Non-Hispanic |
| **participant 33** | no psychotropic<br>medicines | HC | N/A | 34–38 | Male | White | Non-Hispanic |
| **participant 34** | no psychotropic<br>medicines | HC | N/A | 19–23 | Male | Asian | Non-Hispanic |
| **participant 35** | no psychotropic<br>medicines | HC | N/A | 29–33 | Female | White | Non-Hispanic |
| **participant 36** | no psychotropic<br>medicines | HC | N/A | 29–33 | Male | White | Non-Hispanic |
| **participant 37** | no psychotropic<br>medicines | HC | N/A | 29–33 | Male | White | Hispanic |
| **participant 38** | no psychotropic<br>medicines | HC | N/A | 24–28 | Female | White | Non-Hispanic |
| **participant 39** | stimulants | HC | N/A | 29–33 | Female | White | Non-Hispanic |
| **participant 40** | no psychotropic<br>medicines | HC | N/A | 34–38 | Male | White | Non-Hispanic |
| **participant 41** | no psychotropic<br>medicines | HC | N/A | 39–43 | Female | Black or<br>African<br>American | Non-Hispanic |
| **participant 42** | no psychotropic<br>medicines | HC | N/A | 34–38 | Female | White | Non-Hispanic |
| **participant 43** | no psychotropic<br>medicines | HC | N/A | 24–28 | Female | White | Hispanic |
| **participant 44** | no psychotropic<br>medicines | HC | N/A | 24–28 | Female | White | Non-Hispanic |
| **participant 45** | no psychotropic<br>medicines | HC | N/A | 39–43 | Male | White | Hispanic |
| **participant 46** | no psychotropic<br>medicines | HC | N/A | 39–43 | Male | White | Non-Hispanic |
| **participant 47** | no psychotropic<br>medicines | HC | N/A | 29–33 | Female | White | Non-Hispanic |
| **participant 48** | no psychotropic<br>medicines | HC | N/A | 44–48 | Male | White | Non-Hispanic |

**Supplementary Table 2.**
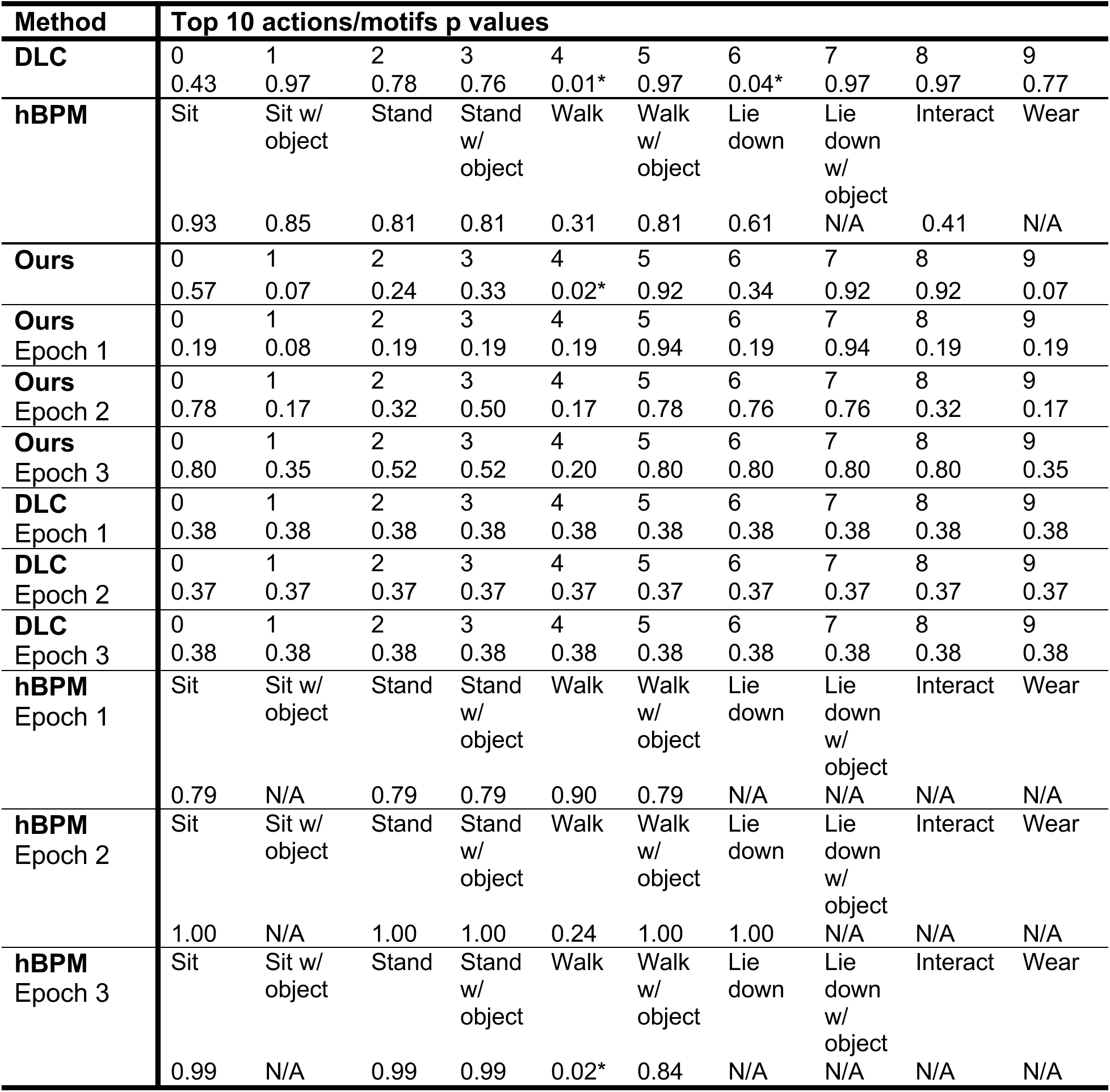
Two sample T-test (Benjamini-Hochberg adjusted p values) dwell time between BD and HC.

**Supplementary Table 3.**
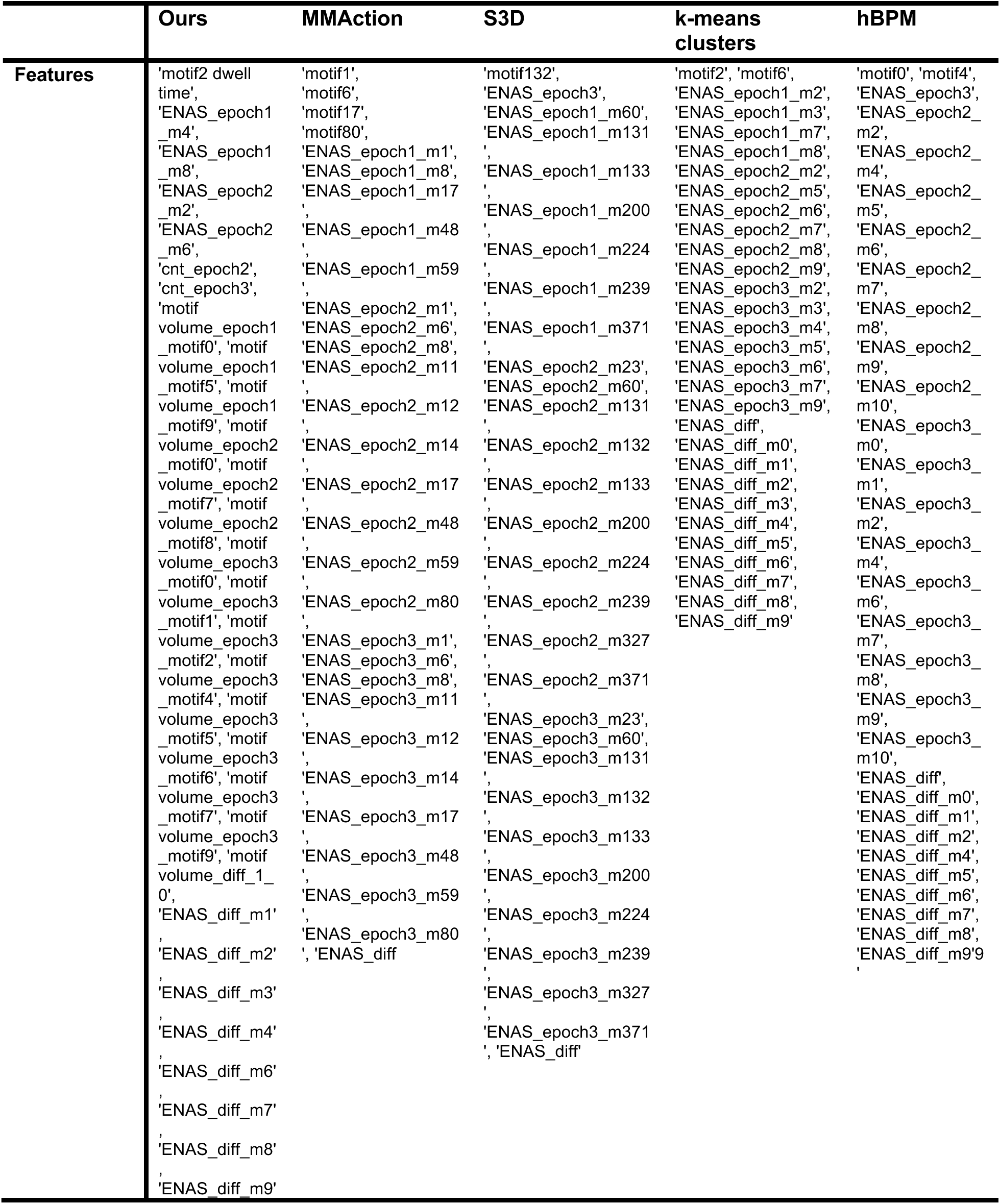
Selected Features in Each Approach.

**Supplementary Table 4.** Pearson Correlation Coefficient r between motif-volume and dwell time (mean ± std)

| **Motif** | **HC** | **BD** |
| --- | --- | --- |
| *Motif 0* | 0.07 ± 0.39 | -0.13 ± 0.38 |
| *Motif 1* | -0.08 ± 0.56 | -0.37 ± 0.48 |
| *Motif 2* | -0.05 ± 0.43 | -0.06 ± 0.35 |
| *Motif 3* | 0.86 ± 0.00 | -0.02 ± 0.12 |
| *Motif 4* | N/A | -0.76 ± 0.05 |
| *Motif 5* | -0.24 ± 0.00 | 0.48 ± 0.00 |
| *Motif 6* | 0.24 ± 0.00 | 0.37 ± 0.20 |
| *Motif 7* | 0.23 ± 0.17 | 0.11 ± 0.02 |
| *Motif 8* | 0.73 ± 0.05 | 0.51 ± 0.10 |
| *Motif 9* | 0.0004 ± 0.3420 | 0.5055 ± 0.0626 |

**Supplementary Table 5.**
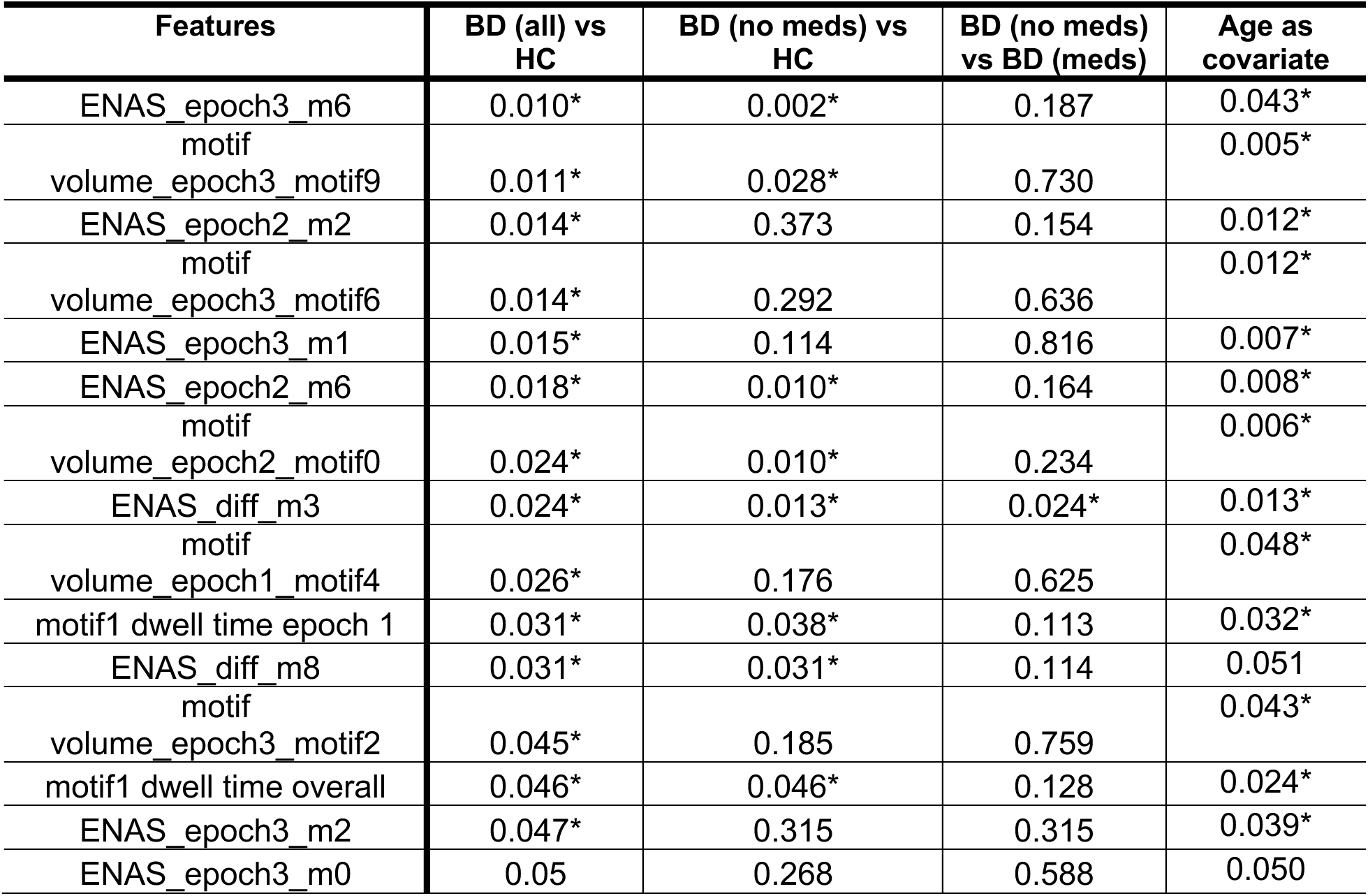
Statistical Test (p-value) for BD (all/no meds) vs HC, BD (no meds) vs BD (meds) in Top Features.

**Supplementary Figure 1.**
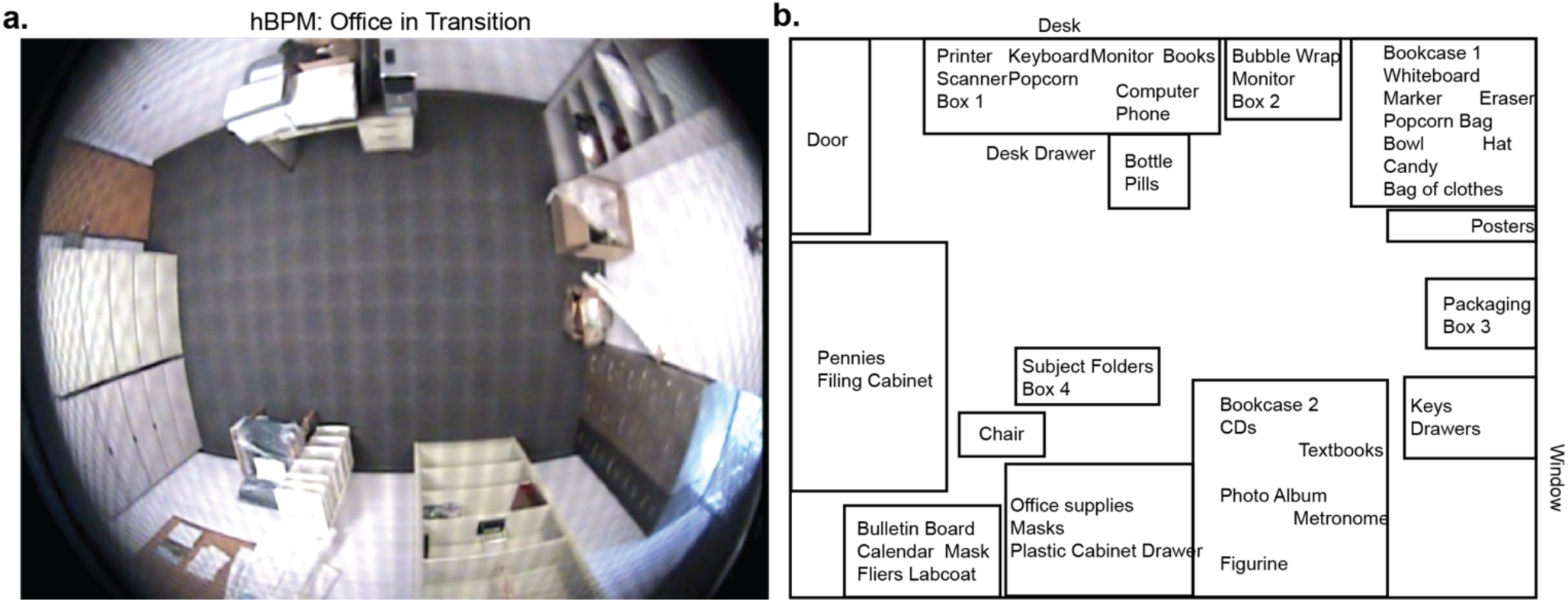
hBPM room setting. **a.** hBPM is an office in transition with objects. **b**. Objects reference map. The bulletin board, the filing cabinet, bookcases 1 and 2, and the desk were placed on the periphery of the room. The chair was placed in a way to prevent sitting.

**Supplementary Figure 2.**
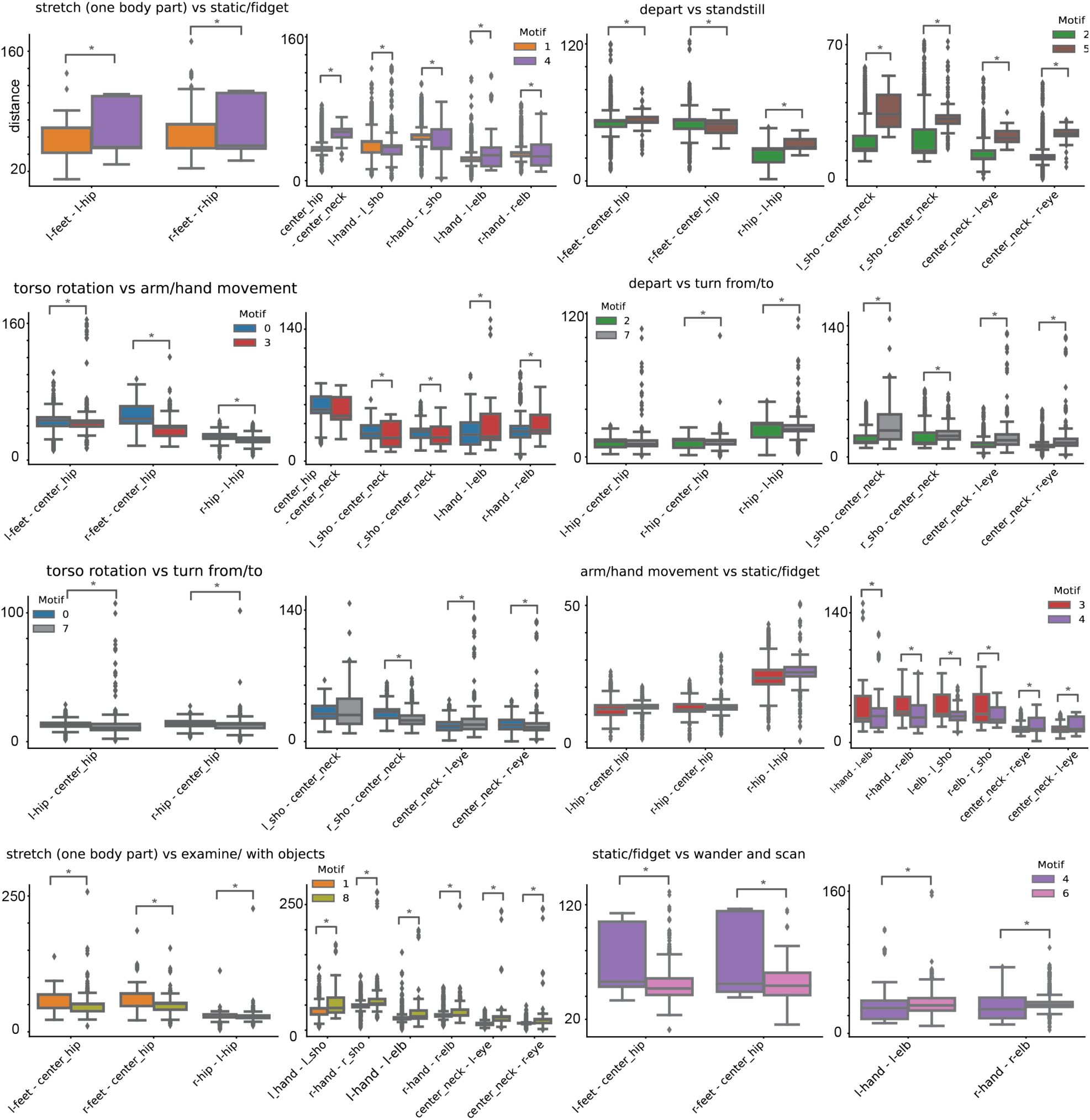
Comparison of pairwise key-point distances across *motifs*. Box plots displaying the distribution of pairwise distances for selected lower body (left) and upper body (right) key-point pairs for example *motifs* pairs. Colors represent different motif pairs being compared. Significance marked by asterisks.

**Supplementary Figure 3.**
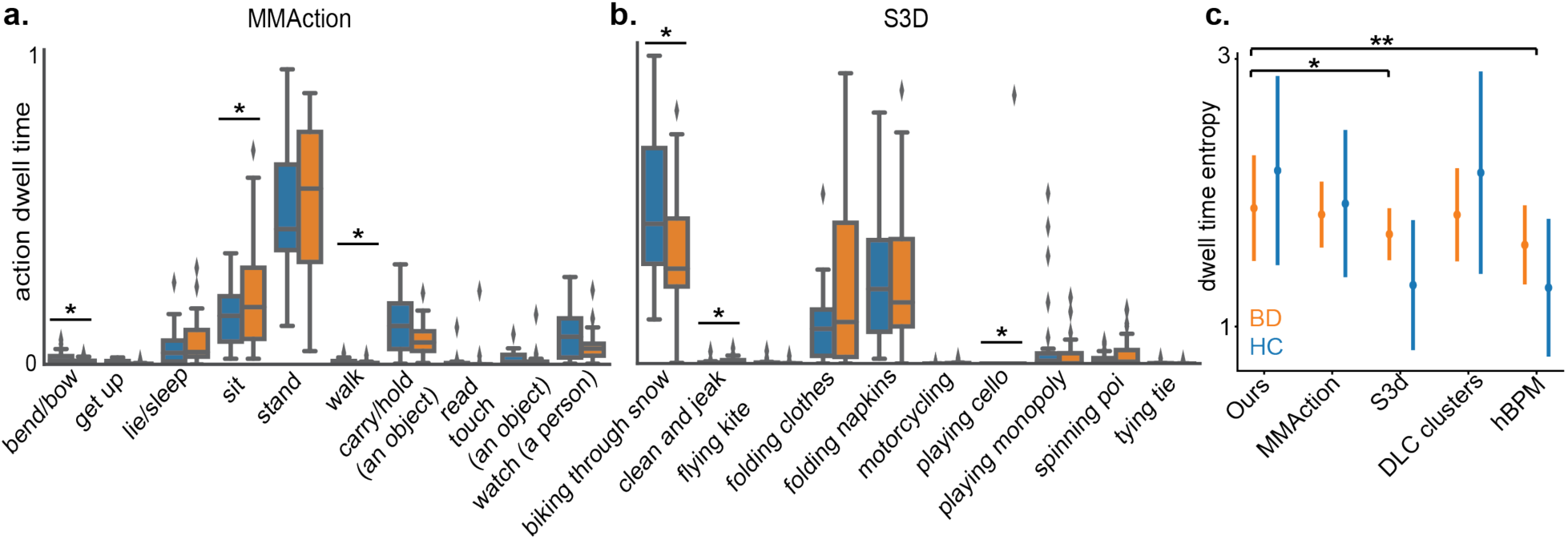
The dwell time of CV-identified actions. **a.** Action dwell time (normalized to a range of [0, 1]) from the top 10 action labels from MMAction in BD (orange) and HC (blue). **b.** Action dwell time (normalized to a range of [0, 1]) from the top 10 action labels from S3D in BD (orange) and HC (blue). **c.** Motif entropy of the dwell time of each approach (mean ± std). Significance marked by asterisks.

**Supplementary Figure 4.**
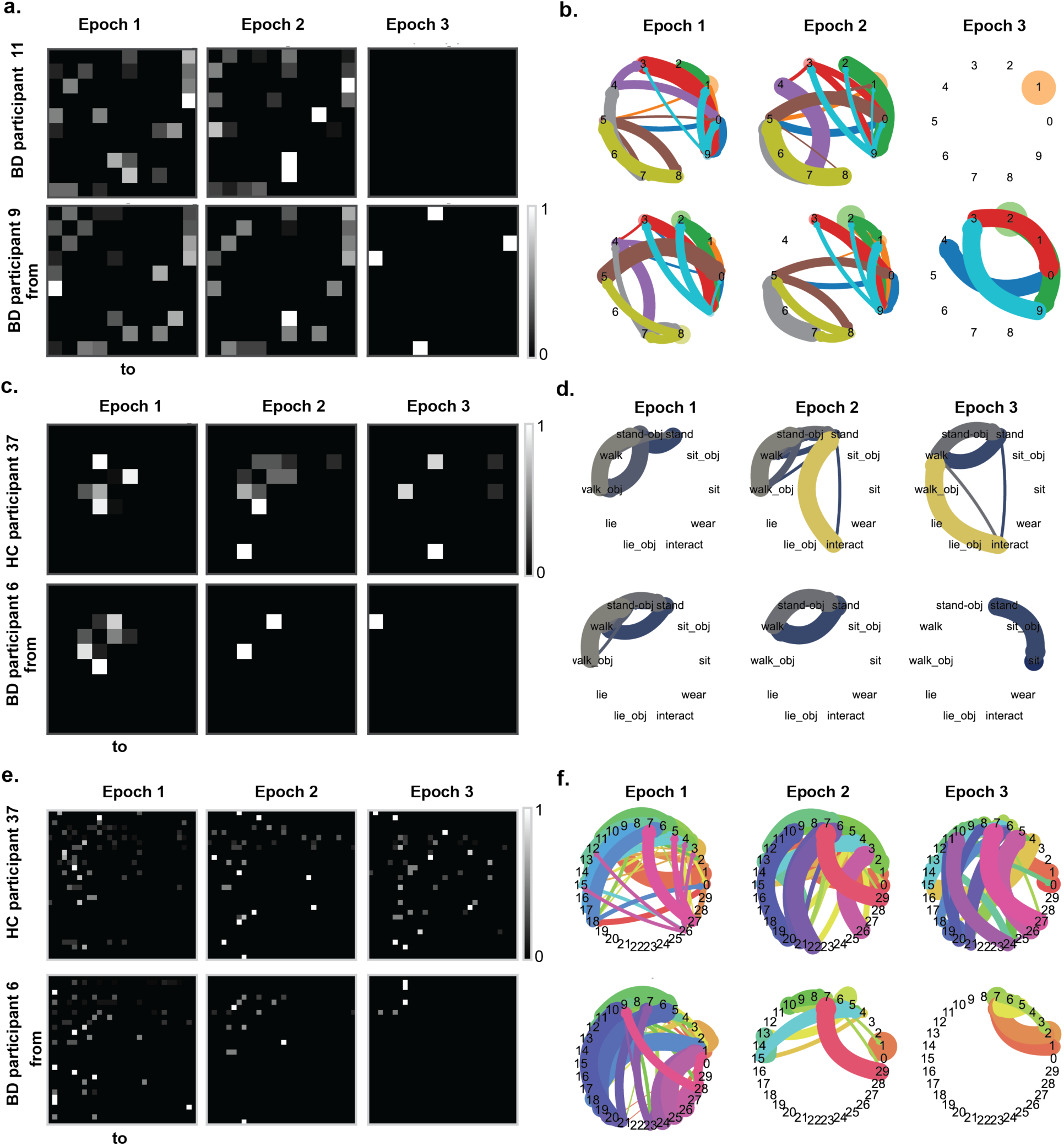
More examples of motif transition, and motif transition in human annotation and control condition. **a.** Examples of transition matrices in three epochs for an additional two individual BD participants (participants 9 and 11) and **b.** graphs representing these transition matrices. Nodes represent motifs and directed edges indicate transition directions and are colored by the ‘from’ motif color. The thicker the edges the higher the transition probability. The larger the nodes the higher the dwell time of the motif. **c.** Examples of human-annotated actions transition matrices in three epochs for the same HC participant 37 and BD participant 6 are shown in Fig.3, **d.** Graphs representing the transition matrices in c. “_obj” refers to an action label with interaction with an object. **e.** Transition matrices in three epochs with n = 30 motifs for the same HC participant 37 and BD participant 6 in Fig.3, where each pixel represents the transition probability from every motif into every other motif. **f.** Graphs representing the transition matrices in e.

**Supplementary Figure 5.**
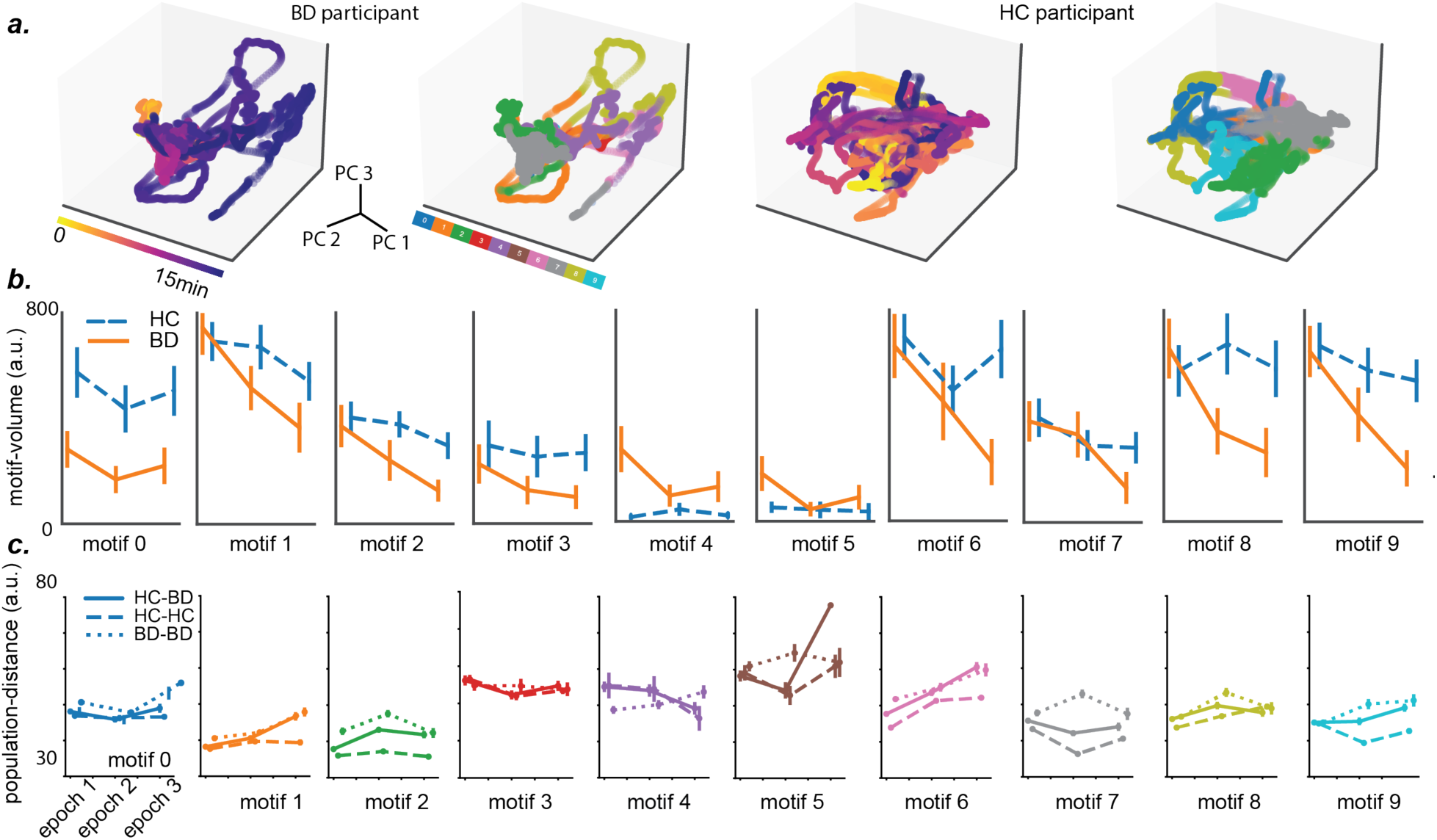
Motif-volume and distance of other *motifs*. **a.** Latent trajectory in top three PC space (left) and latent segmentation of motifs (right) for the same HC participant 37 and BD participant 6 in Fig 3. **b.** *motif-volume* (mean ± std) in BD (orange) and HC (blue) in epochs 1, 2, and 3. **c.** population-distance (mean ± std), including interpopulation-distance (solid lines), intrapopulation-distance of BD-BD (dotted lines) and HC-HC (dashed lines) in epochs 1, 2, and 3.

